# Effects of cognitive training under inspiratory hypoxia on cognition and neuroplasticity in healthy humans: a randomised, double-blind, controlled, four-arm trial

**DOI:** 10.64898/2026.06.28.26356414

**Authors:** Viktoria Damgaard, Johanna Mariegaard Schandorff, Annette Johansen, Julian Macoveanu, Katrine Cramer, Ida Plougmann Østergaard, Katrine Krabbe Thommesen, Caroline Fussing Bruun, Morten Meyer, Pontus Plavén-Sigray, Szabolcs Lehel, Claus Svarer, Gitte Moos Knudsen, Martin Balslev Jørgensen, Lars Vedel Kessing, Hannelore Ehrenreich, Kamilla Woznica Miskowiak

## Abstract

Moderate hypoxia is increasingly recognized as a physiological driver of neuroprotection and neuroregeneration. In this first randomised, double-blind, controlled, four-arm trial, we demonstrate the cognitive and neuroplastic effects of cognitive training under moderate inspiratory hypoxia in humans. Healthy volunteers underwent three weeks of either cognitive or sham training under normobaric hypoxia (12% O_2_) or normoxia (20% O_2_) for 3.5 hours daily, six days per week. Participants were assessed at baseline, treatment completion, and one-month follow-up. The primary outcome was change in a broad cognitive composite score. Additional cognitive, blood-based, and neuroimaging outcomes were assessed, including measurement of the presynaptic protein SV2A with [^11^C]UCB-J positron emission tomography (PET) and neural activity through functional magnetic resonance imaging (fMRI). In total, 126 participants were randomised to hypoxia-cognitive training (H-CT: *n*=36), hypoxia-sham training (H-ST: *n*=30), normoxia- cognitive training (N-CT: *n*=30), or normoxia-sham training (N-ST: *n*=30). Intention-to-treat analyses showed no effect of H-CT relative to N-ST in the primary outcome at treatment completion (primary endpoint; treatment effect=0.11, 95% CI=[-0.06;0.28], *p*=0.19), but improvements emerged at follow-up (treatment effect=0.17, 95% CI=[0.01;0.34], *p*=0.04). N-CT induced transient improvement in the primary outcome at treatment completion (treatment effect=0.20, 95% CI=[0.02;0.38], *p*=0.03), which rendered non-significant at follow-up. Finally, H-ST showed no significant cognitive change relative to N-ST. Moderate hypoxia was safe and well-tolerated. Cognitive benefits were accompanied by decreased hippocampal presynaptic density measured with [^11^C]UCB-J PET. In conclusion, three weeks of H-CT can enhance cognition with associated effects on neuroplasticity, although with a delayed onset of effects on cognition.

## Introduction

Cognitive impairment is a key phenotype of neuropsychiatric disorders that contribute to diminished quality of life, occupational disability, and significant socioeconomic burden [1, 2]. Although cognitive training interventions have demonstrated benefits on cognition across psychiatric populations, effects are generally small-to-moderate and transient [3–5]. Neuroplasticity, including neurogenesis and synaptic reformation in response to internal or external stimuli, play a crucial role in adaptive central nervous system (CNS) function and cognitive health, and disruption in psychiatric disorders may give rise to cognitive deficits [6]. Given the limited effectiveness of cognitive training alone, multimodal interventions in which cognitive training is complimented by additional strategies that directly target and restore neuroplasticity pathways may be necessary to produce *sustained* cognitive improvement [7].

Hypoxia - a reduction in cellular or tissue oxygen levels relative to physiological norms - is emerging as a therapeutic strategy. Contrary to the common perception of hypoxia as solely detrimental, accumulating evidence indicates that hypoxia in adequate doses can target various CNS pathways that improve neuroplastic potential [8–11]. In a systematic review, we found that moderate hypoxia interventions across humans and animals involving prolonged (>two weeks) repeated exposure to 10-16% O_2_ induced beneficial effects on cognition and markers of neuroplasticity [12]. The mechanisms involve a transcriptional program that is regulated in part by hypoxia-inducible factors (HIFs), which modulate gene expression in response to reduced cellular oxygen availability, alongside other HIF-independent mechanisms [13–18]. Among the genes regulated by HIFs are several trophic and growth factors, such as erythropoietin (EPO) and vascular endothelial growth factor (VEGF), that promote neuroplasticity and serve as endogenous neuroprotective mechanisms [15, 18–20]. In particular, exogenously administered EPO has experimentally been shown to improve aspects of cognition in healthy individuals [21], as well as in people with schizophrenia [22], bipolar disorder [23], depression [24], and multiple sclerosis [25] - effects that were accompanied by increase in aberrant task-related dorsolateral prefrontal cortex (DLPFC) activity [26, 27]. Supporting a role of EPO in hypoxia-induced neuroplasticity, previous studies in mice demonstrated that three weeks of 12% O_2_ combined with motor-cognitive training enhances complex learning and hippocampal plasticity, mediated by upregulation of brain EPO and its receptor (EPOR) [10].

Despite these encouraging findings, animal models of CNS conditions have limited predictive value for treatment efficacy in humans [28]. While preliminary results from pilot studies (*N*<18 per group) indicated positive effects of hypoxia combined with physical exercise interventions in geriatric populations, no randomised controlled trials have yet evaluated the efficacy of concurrent cognitive training and hypoxia in humans [12]. Therefore, this is the first double-blinded, randomised, controlled study to investigate the effects of repeated normobaric inspiratory hypoxia (12% O_2_) with concurrent cognitive or sham training compared with normoxia (20% O_2_) with cognitive or sham training in healthy humans. Building on previous studies in mice [10], the present trial involved 3.5 hours daily hypoxia/normoxia exposures with concurrent cognitive/sham training six days per week over three weeks. We hypothesized that hypoxia with cognitive training (H-CT) would lead to (i) immediate increase in a broad cognitive composite measure, ‘speed of complex cognitive processing’, (primary outcome) that would be sustained at the one-month follow-up assessment relative to normoxia with sham training (N-ST), and (ii) that cognitive improvements would be accompanied by change in presynaptic density in the hippocampus and the frontal cortex and increase in task-related DLPFC activation (mimicking the neural effects of exogeneous EPO treatment [26, 27, 29]). We also evaluated the separate effects of hypoxia with sham training (H-ST) and cognitive training under normoxia (N-CT). Here, we hypothesized that (iii) each monotherapy would yield immediate, but transient, cognitive effects that would have disappeared one month post-treatment relative to N-ST.

## Materials and methods

### Study design and participants

The trial had a randomised, controlled, double-blinded, parallel-group design and was conducted in parallel with a smaller open-label clinical trial investigating the effects of combined H-CT compared to waitlist in affective disorders (open access study protocol: [30]). Data were collected across three sites: Department of Psychology, University of Copenhagen, Psychiatric Centre Copenhagen, Frederiksberg Hospital, and Copenhagen University Hospital, Rigshospitalet. Participants were recruited from education facilities in Copenhagen, Denmark and online advertisements. All participants were between 18 and 50 years, fluent in Danish, and somatically and psychiatrically healthy (details in **Supplement**). After inclusion of the 120 participants (see power calculation below), we enrolled an extra group of six (also blinded) participants allocated to the H-CT group to obtain a sufficient sample size for the positron emission tomography (PET) imaging analysis, because the PET scanner had been temporarily unavailable, and more participants were thus needed for sufficient statistical power (details in **Supplement**).

The study was approved by the Health Research Ethics Committee in the Capital Region of Denmark (H-22028111) and Danish Data Protection Agency (P-2022-354) and preregistered at ClinicalTrials.gov (NCT06121206). The CONSORT 2025 reporting guidelines were followed [31]. All participants were informed about the study procedures, and written informed consent was obtained in accordance with the Declaration of Helsinki. Patient and public involvement was not undertaken in the design or reporting of this study due to its mechanistic aim.

### Randomisation and blinding

Participants were randomised using a 1:1 allocation ratio in an automated randomisation module in the online Research Electronic Data Capture (REDCap) system based on a blocked randomisation list created by an independent researcher through Sealed Envelope Ltd. [32]. Randomisation was conducted after baseline assessments, with participants scheduled to undergo treatment together in groups of two-four. The prespecified age-based stratification (<27 or ≥27 years) could not be implemented, as participants who volunteered to undergo treatment sessions together were typically not from the same age stratum. However, the groups were well balanced with respect to age (**Table 1**). The randomisation list was inaccessible to the researcher who enrolled and randomised participants to prevent unblinding. Participants and outcome assessors were blinded to both oxygen and training conditions, and participants were instructed not to talk about their treatment during treatment sessions and outcome assessments.

**Table 1:** Baseline demographic characteristics in the four treatment groups.

|  | **H-CT** | **H-ST** | **N-CT** | **N-ST** |
| --- | --- | --- | --- | --- |
| *N* | 36 | 30 | 30 | 30 |
| Age, years | 25.0 (23.0-27.5) | 25.0 (24.0-28.0) | 26.0 (22.0-30.0) | 25.0 (22.0-29.0) |
| Sex, no. female (%) | 20/36 (56%) | 15/30 (50%) | 15/30 (50%) | 15/30 (50%) |
| Verbal IQ | 108.9 (106.4-111.0) | 108.9 (106.4-110.6) | 109.7 (107.3-113.1) | 108.5 (106.4-110.6) |
| Years of education | 15.0 (13.0-17.0) | 15.0 (13.0-17.0) | 15.0 (13.0-17.0) | 14.0 (13.0-16.0) |
| Employment (full or part time), no. yes (%) | 12/36 (33%) | 6/30 (20%) | 7/30 (23%) | 11/30 (37%) |
| Student, no. yes (%) | 22/36 (61%) | 19/30 (63%) | 17/30 (57%) | 17/30 (57%) |
| **Note:** Continuous values are shown as “n (n-n)” (indicating median and its 25th and 75th percentiles). **Abbreviations:** H-CT=Hypoxia with cognitive training; H-ST=Hypoxia with sham training; IQ=Intelligence quotient; N-CT=Normoxia with cognitive training; N-ST=Normoxia with sham training. |  |  |  |  |

### Procedure

Participants were randomised to one of four arms: I) hypoxia (12% O_2_) with cognitive training (H-CT), II) hypoxia with sham training (H-ST), III) normoxia (20% O_2_) with cognitive training (N-CT), or IV) normoxia with sham training (N-ST). Treatment sessions were 3.5 hours, repeated six days per week over three weeks. Cognitive/sham training was conducted concurrently with hypoxia/normoxia exposure for feasibility reasons. Prior to inclusion in the study, participants underwent a comprehensive screening to ensure compliance with inclusion and safety criteria. The baseline assessment was conducted the week prior to beginning the treatment (week 0) by a psychologist or trained psychology student. At baseline, an estimate of a verbal intelligence quotient (IQ) was further obtained using the Danish Adult Reading Test (DART). Assessments were repeated at treatment completion (week 4; primary endpoint) and at a one-month follow-up (week 8). PET scans with the radioligand [^11^C]UCB-J that binds to the synaptic vesicle glycoprotein 2A (SV2A) as a readout of presynaptic density were carried out in week 4 for participants in the two extreme groups (i.e., H-CT and N-ST). Participants were also assessed with fMRI at baseline and week 8. Follow-up fMRI was performed in week 8 to allow normalization of expected hypoxia-related change in red blood cell parameters that could otherwise confound the blood-oxygen-level-dependent (BOLD) signal. Haematological tolerability parameters (haemoglobin, erythrocytes, reticulocytes, and thrombocytes values) were measured at baseline, prior to beginning days 8 and 19 of treatment, and week 8. Blood samples for analysis of serum EPO, VEGF, and brain-derived neurotrophic factor (BDNF) were collected at baseline and immediately prior to day 19 (more details in the **Supplement**).

### Treatment groups

#### Hypoxia and normoxia

Normobaric air with either 12% or 20% O_2_ were blown into a sealed 20 m^3^ treatment room by a 4kW air compressor with a safety-approved system developed HöhenBalance, Austria. For the hypoxic condition, participants entered the room at 16% O_2_. On the first treatment day, oxygen levels were gradually lowered from 16% to 12% O_2_ (approx. 4,400 meters altitude) over a two-hour lead-in phase to minimize risk of side effects. If no adverse effects were observed, oxygen levels were reduced from 16% to 12% O_2_ over *Mdn*=46 minutes (IQR 43-52) in the subsequent sessions. For the normoxic condition, the oxygen level wat set at 20% O_2_. This small reduction from 21% O_2_ (sea level) was chosen to ensure blinding due to humming noises from the compressor located outside the building. The room was also of equal temperature and humidity in both conditions to maintain blinding. All participants wore a pulse oximeter (Shanghai Berry Electronic Tech Co., Ltd) that measured SpO_2_ and pulse rate during sessions, which were continuously monitored by unblinded research personnel. Altitude-related cerebral side effects were systematically assessed after each session with the Environmental Symptoms Questionnaire (ESQ) [33]. In the event of altitude sickness or other medical emergencies, medical doctors were available. Adverse events (AEs) were monitored continuously, and any serious AEs were reported to the local ethical committee.

#### Cognitive and sham training

The cognitive training was provided by the web-based program, Happy Neuron Pro (Danish version, 2024), which has previously shown pro-cognitive effects [34, 35]. The program builds upon principles of neuroplasticity-based learning by being intensive, adaptive, and rewarding. It consisted of 26 different exercises, each with 30 difficulty levels that increased following consecutive trials with 80% accuracy. The sham training involved similar exercises but with low cognitive demand that has previously shown to produce no cognitive benefits [36]. The sham training included the exact same stimuli as the cognitive training condition, but with no adjustments from trial to trial except in the appearance of the exercises. All participants were instructed to train for two hours per session, interleaved with short breaks inside the altitude room where participants relaxed or slowly walked on a treadmill. More details on the interventions are available in the **Supplement**.

### Outcome measures

The predefined primary outcome was change in a cognitive composite score, ‘speed of complex cognitive processing’, from baseline to treatment completion (week 4). This was based on our prior trial demonstrating improvement in this measure after exogenous EPO treatment [23]. The composite score was computed by *z*-standardising raw test scores based on the baseline means and standard deviations (SD) across all participants, and then averaging *z*-scores across the following tests of attention, processing speed, verbal memory, and executive functions: Rey Auditory Verbal Learning Test (RAVLT) lists I-V total recall, Repeatable Battery for the Assessment of Neuropsychological Status (RBANS) Coding, Verbal Fluency (letter “D”), Wechsler Adult Intelligence Scale (WAIS)-III Letter-Number Sequencing, Trail Making Test Part B, and Rapid Visual Information Processing (RVP) from CANTAB (Cambridge Cognition Ltd.). To minimize learning effects from repeated testing, matched alternate versions of the RAVLT and RBANS Coding were used in a counter-balanced order.

The co-secondary cognitive outcome was change in ‘mean choices to correct’ in the One Touch Stockings of Cambridge (OTS) task from CANTAB, based on our previous trial showing improvement in this measure after similar cognitive training [35]. The co-secondary neural outcome was change in DLPFC activity during a spatial working memory N-back fMRI task. This area has previously shown target engagement in response to pro-cognitive interventions, including exogenous EPO treatment, in both healthy and neuropsychiatric populations, and thus present as a putative biomarker for pro-cognitive effects (e.g., [26, 37–40]).

Tertiary cognitive outcomes were change in global and domain-specific performance based on the full neuropsychological test battery. Tertiary self-report outcomes were domains of subjective cognition, quality of life, sleep, and functioning. Mechanistic outcomes included [^11^C]UCB-J non-displaceable binding potential (*BP*_ND_) in the hippocampus (key hub for neurogenesis) and frontal cortex (involved in general cognitive proficiency) at treatment completion, change from baseline to follow-up in task-related neural activation across the whole dorsal prefrontal cortex, and change in serum EPO, VEGF, and BDNF from baseline to day 19 of treatment. For full list of outcomes and calculations, see the **Supplement**.

### PET analysis

Details on PET acquisition, image preprocessing, kinetic modelling, and missing data are available in the **Supplement**. In brief, participants underwent a six min transmission scan followed by a 90 min emission scan which started at the time of the intravenous [^11^C]UCB-J bolus injection administered over 20 seconds. Motion correction was applied using the Automated Image Registration (AIR) software with the reconcile command (v. 5.2.5) [41]. PET images were co-registered to the participant’s T1-weighted MR image from their follow-up scan, and tissue time-activity curves were extracted from automatically defined regions of interest (ROIs) [42–44] using the PVElab software pipeline (https://nru.dk/pveout/). Quantification was performed using a simplified reference tissue model 2 (SRTM2) to estimate *BP*_ND_ with centrum semiovale (white matter) as reference region [45]. Our a priori measures of interest were the hippocampus and frontal cortex, however, we also extracted estimated *BP*_ND_ for other regions to explore eventual group differences in presynaptic density across the whole brain (**Supplement**).

### fMRI analysis

Details on the visuospatial working memory N-back paradigm, fMRI data acquisition, preprocessing and analysis, missing data, and in-scanner behaviour data analysis are available in the **Supplement**. Briefly, the fMRI data were pre-processed using fMRIPrep (v. 25.1.1) [46]. Subject- and group-level analyses were conducted in FMRI Expert Analysis Tool (FEAT) (v. 6), part of FMRIB’s Software Library (FSL) [47]. At the subject level, the task was modelled using two events: 2-back>0-back (general working memory) and 2-back>1-back (high-load specific working memory) that were convolved with a double-gamma hemodynamic response function [48]. We constructed an 8 mm spherical ROI around the right DLPFC (x=40, y=34, z=29) given the paradigm’s visuospatial component and primary engagement of the right hemisphere, consistent with our prior studies [27, 38, 40]. The mean percent BOLD signal change for the DLPFC ROI was extracted using the featquery tool for general working memory (co-secondary outcome) and high-load working memory (exploratory) for subsequent statistical analysis. To explore any unpredicted treatment-related neural activity changes in the dorsal prefrontal cortex and across the whole brain, we conducted separate two-way mixed effects ANOVAs in FEAT with complete-cases only. The models used FMRIB’s Local Analysis of Mixed Effects (FLAME) as estimation method [49] with *p*<0.05, and corrected for multiple comparisons with a cluster-forming threshold of *Z*=2.56 (*p*<0.005).

### Statistical analysis

A power calculation performed using data from our previous clinical study of exogenous EPO treatment [23] showed that 104 participants (26 per arm) would achieve >80% power to detect a greater improvement of 0.40 or more *z*-scores (medium effect size) in the H-CT vs. N-ST groups for the primary outcome (hypothesis i) at an α-level of 0.05 [30]. To accommodate an expected 20% attrition rate, we included 30 participants per treatment arm.

Statistical analyses were performed in R (v. 2026.01.0). Variable distributions were visually inspected and formally assessed with Shapiro-Wilk tests. All individual cognitive and self-report outcome measures were *z*-standardized and inversed where appropriate, so a higher score reflected better performance. All participants with baseline data were included in the intention-to-treat analyses. To investigate the longitudinal treatment effects on our predefined primary, secondary, and tertiary outcomes, we conducted linear mixed models (LMMs) with an unstructured covariance pattern. Fixed effects were time (baseline, treatment completion, and follow-up), treatment (H-CT vs. H-ST vs. N-CT vs. N-ST), and their interaction (time*treatment). N-ST was entered as the reference group, and baseline constraint was applied. For the primary outcome, the significance level was set to *p*<0.05 (two-tailed). The BH method was applied to adjust for multiple comparisons separately for secondary and tertiary outcomes. The false discovery rate (FDR) was set at 5%, and adjusted *p*-values<0.05 were considered significant.

For outcomes showing a significant effect of H-CT (vs. N-ST), we performed separate exploratory LMMs to assess potential synergistic effects between the two interventions (hypoxia*cognitive training*time). Blood-based tolerability and biomarker measures were analysed with LMMs similar to those used for the outcome analyses. To examine group differences in presynaptic density at treatment completion, we compared average [^11^C]UCB-J *BP*_ND_ between H-CT and N-ST groups with independent samples *t*-tests. Finally, group comparisons in treatment fidelity and tolerability measures were tested with one-way analyses of variance (ANOVAs) with Tukey’s post hoc tests or Kruskal–Wallis tests with Benjamini-Hochberg (BH)-adjusted pairwise comparisons (all four groups) and independent samples *t*-tests or Mann–Whitney *U* tests (hypoxia vs. normoxia groups). Further details are available in the **Supplement**.

## Results

### Participant flow and missing data

See **Figure 1** for the CONSORT flow diagram and exclusion/discontinuation reasons. A total of 152 individuals were screened for eligibility, of whom 126 participants were enrolled and randomised between February 2023 and June 2025 (H-CT: *n*=36; H-ST: *n*=30; N-CT: *n*=30; N-ST: *n*=30). Twelve participants (10%) withdrew from the study before treatment completion. Of these, four (3%) withdrew prior to commencing treatment, while eight (7%) withdrew during treatment. No participants were lost to one-month follow-up. The treatment groups were well-balanced (**Table 1**). Data were analysed for all 126 participants with baseline data as per the intention-to-treat principle. For the PET imaging analysis, a subsample of 30 well-matched participants (H-CT: *n*=16, N-ST: *n*=14) were analysed (more details and flow diagram are available in the **Supplement**).

**Figure 1.**
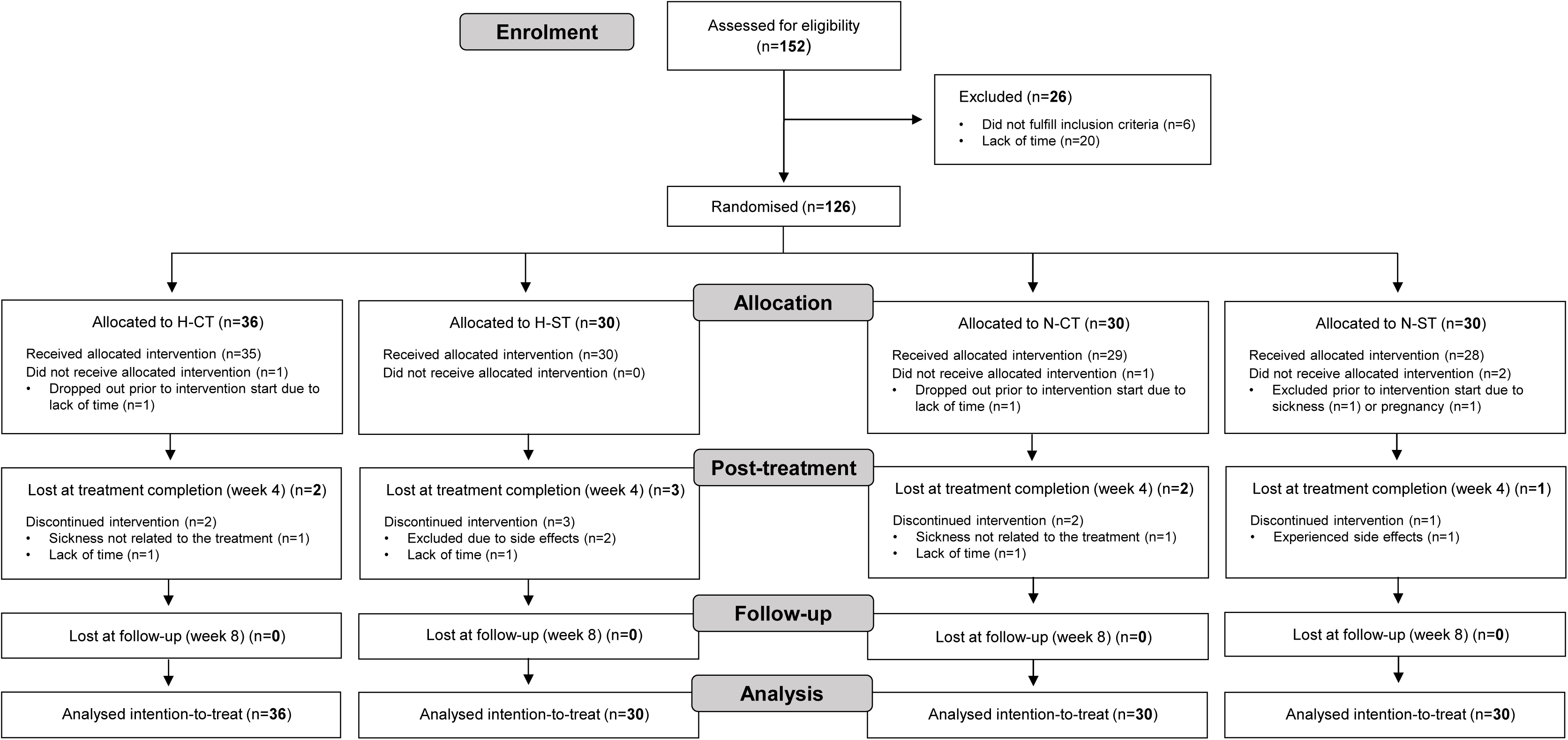
CONSORT flow diagram.

### Primary and secondary outcomes

#### H-CT vs. N-ST

There was no effect of combined H-CT (vs. N-ST) on the primary outcome, ‘speed of complex cognitive processing’, at treatment completion (primary endpoint; treatment effect=0.11, 95% CI=[-0.06;0.28], *p*=0.19). However, H-CT-related improvements emerged at one-month follow-up (treatment effect=0.17, 95% CI=[0.01;0.34], *p*=0.04) (**Table 2**; **Figure 2**). This finding prevailed in a post hoc sensitivity analysis removing the six additional participants in the H-CT group (*p*=0.03). Exploratory LMM analysis showed no showed no statistically detectable synergistic effects between hypoxia and cognitive training in the primary outcome at follow-up (*p*=0.63). There were no effects of H-CT (vs. N-ST) on the co-secondary outcomes OTS mean choices to correct (*ps*≥0.56) or right working memory-related DLPFC activity (*p*=0.47) (**Table 2**).

**Figure 2.**
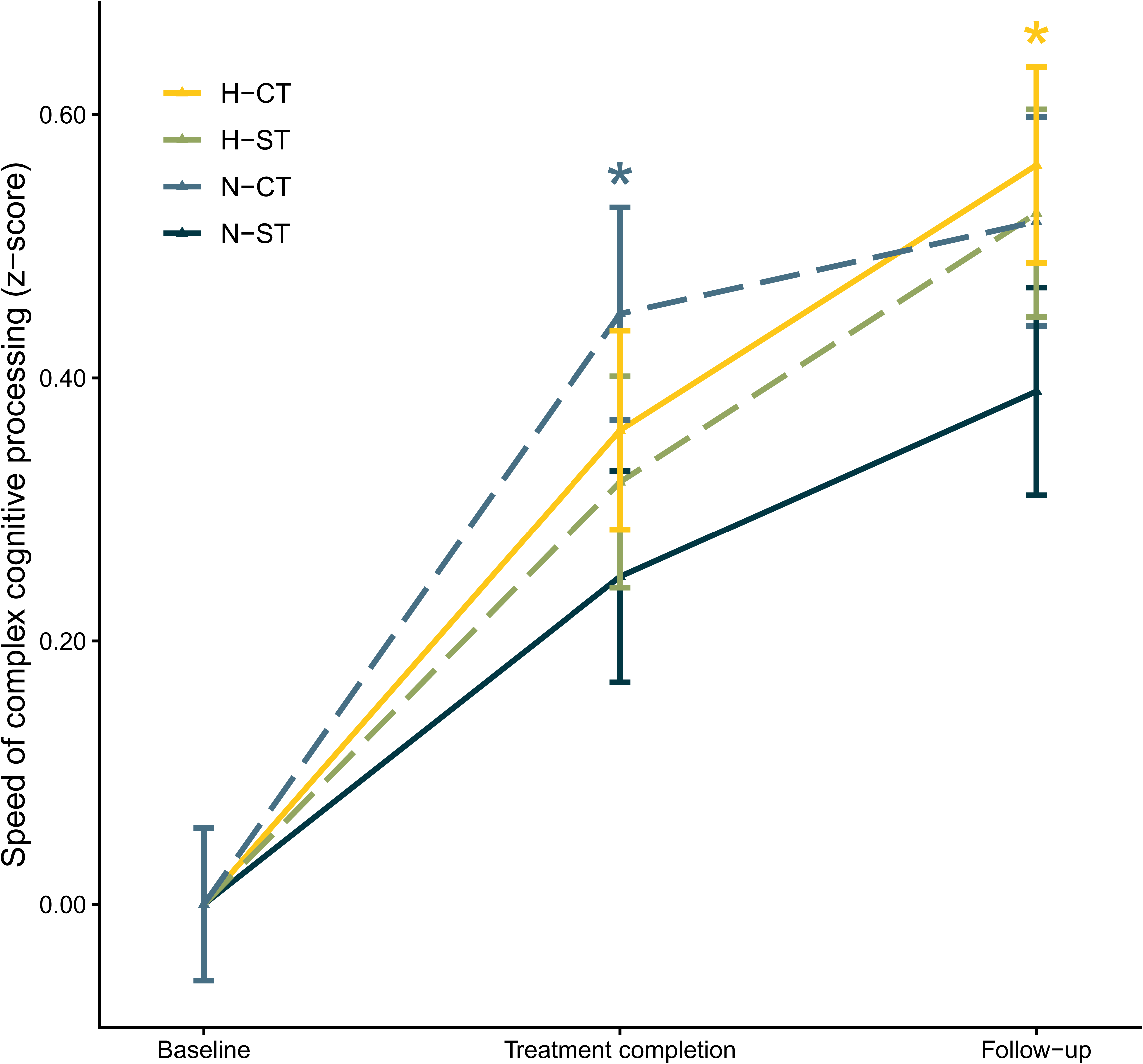
Effects of hypoxia and/or cognitive training on the primary outcome, a cognitive composite score of ‘speed of complex processing’, at treatment completion (week 4; primary endpoint) and one-month follow-up (week 8). **Legend:** There was no effect of combined H-CT relative to N-ST on the primary outcome at treatment completion (primary endpoint), but statistically significant modest effects emerged at one month post-treatment. In contrast, N-CT produced immediate improvements at treatment completion, which did not prevail at follow-up. H-ST elicited no significant change relative to N-ST. The y axis shows the predicted effect size from the linear mixed model (in *z*-scores). Error bars show the standard error. \**p*<0.05 relative to N-ST. Abbreviations: H-CT=Hypoxia with cognitive training; H-ST=Hypoxia with sham training; N-CT=Normoxia with cognitive training; N-ST=Normoxia with sham training.

**Table 2:** Effects of hypoxia and/or cognitive training versus normoxia sham training on the primary and secondary predefined outcomes.

|  |  | Treatment completion (week 4) |  |  | Follow-up (week 8) |  |  |
| --- | --- | --- | --- | --- | --- | --- | --- |
|  | Group | Treatment effect | 95% CI | *p*-value | Treatment effect | 95% CI | *p*-value |
| **Primary outcome** |  |  |  |  |  |  |  |
| Speed of complex cognitive processing | H-CT | 0.11 | -0.06; 0.28 | 0.19 | 0.17 | 0.01; 0.34 | **0.04** |
|  | H-ST | 0.07 | -0.11; 0.25 | 0.42 | 0.14 | -0.04; 0.31 | 0.12 |
|  | N-CT | 0.20 | 0.02; 0.38 | **0.03** | 0.13 | -0.04; 0.30 | 0.14 |
|  | N-ST | - | - | - | - | - | - |
| **Secondary cognitive outcome** |  |  |  |  |  |  |  |
| OTS Mean choices to correct | H-CT | 0.03 | -0.28; 0.33 | 0.87 | 0.10 | -0.23; 0.42 | 0.56 |
|  | H-ST | 0.07 | -0.26; 0.40 | 0.66 | 0.08 | -0.27; 0.43 | 0.63 |
|  | N-CT | 0.17 | -0.15; 0.50 | 0.29 | 0.01 | -0.33; 0.36 | 0.95 |
|  | N-ST | - | - | - | - | - | - |
| **Secondary neuroimaging outcome** |  |  |  |  |  |  |  |
| Mean percent BOLD signal change in dorsal prefrontal activity during general working memory (2>0-back)<sup>1</sup> | H-CT | - | - | - | -0.18 | -0.69; 0.32 | 0.47 |
|  | H-ST | - | - | - | -0.14 | -0.67; 0.39 | 0.60 |
|  | N-CT | - | - | - | -0.35 | -0.87; 0.18 | 0.19 |
|  | N-ST | - | - | - | - | - | - |
| **Note:** Outcomes are *z*-standardized based on the baseline means and standard deviations across the whole sample. Includes all available data (incl. study dropouts). <sup>1</sup>Missing data/data not analyzed at baseline for H-ST: *n*=3, N-CT: *n*=1, and N-ST: *n*=4; missing data/data not analyzed at follow-up for H-CT: *n*=2, H-ST: *n*=1, and N-ST: *n*=1. **Abbreviations:** BOLD=Blood oxygen level dependent; CI=Confidence interval; H-CT=Hypoxia with cognitive training; H-ST=Hypoxia with sham training; N-CT=Normoxia with cognitive training; N-ST=Normoxia with sham training; OTS=One Touch Stockings of Cambridge. |  |  |  |  |  |  |  |

#### H-ST and N-CT vs. N-ST

There was an immediate effect of N-CT (vs. N-ST) on ‘speed of complex cognitive processing’ at treatment completion (treatment effect=0.20, 95% CI=[0.02;0.38], *p*=0.03) that did not prevail at follow-up (*p*=0.14) (**Table 2**). H-ST elicited no statistically significant change in this outcome (*ps*≥0.12). There were no effects of N-CT or H-ST (vs. N-ST) on OTS mean choices to correct (*ps*≥0.29) or right DLPFC activation (*ps*≥0.19).

### Tertiary outcomes

Results for the domain-specific tertiary outcomes are shown in **Table 3**. There was a positive effect of H-CT (vs. N-ST) on ‘global cognition’ at treatment completion (*p*=0.03) that prevailed at follow-up (*p*=0.03). There was also a beneficial effect of H-CT on ‘attention’ at follow-up (*p*=0.03) that had not emerged at treatment completion (*p*=0.21). These findings remained significant after removing the six additional H-CT participants in post hoc sensitivity analyses (*ps*≤0.03), except for the improvement in global cognition at treatment completion (*p*=0.054). None of these showed statistically detectable synergistic effects between hypoxia and cognitive training (*ps*≥0.24).

**Table 3:** Effects of hypoxia and/or cognitive training versus normoxia sham training on the tertiary cognitive domain and self-report outcomes.

|  |  | Treatment completion (week 4) |  |  | Follow-up (week 8) |  |  |
| --- | --- | --- | --- | --- | --- | --- | --- |
|  | Group | Treatment effect | 95% CI | *p*-value (adj.) | Treatment effect | 95% CI | *p*-value (adj.) |
| **Cognitive domains** |  |  |  |  |  |  |  |
| Processing speed | H-CT | 0.25 | -0.01; 0.51 | 0.06 | 0.03 | -0.22; 0.29 | 0.81 |
|  | H-ST | 0.16 | -0.11; 0.43 | 0.24 | -0.05 | -0.31; 0.22 | 0.74 |
|  | N-CT | 0.29 | 0.02; 0.56 | **0.03** (0.18) | 0.12 | -0.15; 0.39 | 0.38 |
|  | N-ST | - | - | - | - | - | - |
| Attention | H-CT | 0.12 | -0.07; 0.31 | 0.21 | 0.22 | 0.02; 0.42 | **0.03** (0.26) |
|  | H-ST | 0.09 | -0.11; 0.29 | 0.38 | 0.09 | -0.12; 0.31 | 0.38 |
|  | N-CT | 0.06 | -0.14; 0.26 | 0.55 | 0.11 | -0.11; 0.32 | 0.32 |
|  | N-ST | - | - | - | - | - | - |
| Working memory | H-CT | 0.08 | -0.17; 0.33 | 0.53 | 0.19 | -0.06; 0.45 | 0.13 |
|  | H-ST | 0.12 | -0.14; 0.38 | 0.37 | 0.15 | -0.11; 0.42 | 0.26 |
|  | N-CT | 0.25 | -0.02; 0.51 | 0.07 | 0.15 | -0.11; 0.41 | 0.26 |
|  | N-ST | - | - | - | - | - | - |
| Verbal learning and memory | H-CT | 0.20 | -0.09; 0.50 | 0.18 | 0.20 | -0.16; 0.55 | 0.27 |
|  | H-ST | 0.15 | -0.16; 0.46 | 0.34 | 0.11 | -0.26; 0.48 | 0.55 |
|  | N-CT | 0.34 | 0.03; 0.65 | **0.03** (0.18) | 0.37 | 0.001; 0.74 | **0.049** (0.23) |
|  | N-ST | - | - | - | - | - | - |
| Executive functions | H-CT | 0.15 | -0.04; 0.34 | 0.12 | 0.14 | -0.03; 0.31 | 0.11 |
|  | H-ST | 0.04 | -0.16; 0.24 | 0.67 | 0.07 | -0.11; 0.25 | 0.44 |
|  | N-CT | 0.18 | -0.02; 0.38 | 0.08 | 0.13 | -0.05; 0.31 | 0.16 |
|  | N-ST | - | - | - | - | - | - |
| Global cognition | H-CT | 0.14 | 0.01; 0.27 | **0.03** (0.26) | 0.14 | 0.01; 0.26 | **0.03** (0.26) |
|  | H-ST | 0.09 | -0.05; 0.22 | 0.21 | 0.05 | -0.09; 0.18 | 0.45 |
|  | N-CT | 0.19 | 0.06; 0.32 | **0.005** (0.11) | 0.15 | 0.01; 0.28 | **0.03** (0.18) |
|  | N-ST | - | - | - | - | - | - |
| Facial emotion recognition<sup>1</sup> | H-CT | -0.16 | -0.39; 0.07 | 0.18 | -0.11 | -0.37; 0.14 | 0.39 |
|  | H-ST | 0.04 | -0.20; 0.28 | 0.75 | 0.11 | -0.16; 0.37 | 0.42 |
|  | N-CT | 0.05 | -0.20; 0.30 | 0.70 | 0.01 | -0.27; 0.28 | 0.97 |
|  | N-ST | - | - | - | - | - | - |
| CAVIR global composite | H-CT | - | - | - | -0.04 | -0.28; 0.20 | 0.73 |
|  | H-ST | - | - | - | 0.09 | -0.16; 0.34 | 0.48 |
|  | N-CT | - | - | - | 0.11 | -0.14; 0.36 | 0.38 |
|  | N-ST | - | - | - | - | - | - |
| **Self-report domains** |  |  |  |  |  |  |  |
| Quality of life | H-CT | -0.15 | -0.40; 0.10 | 0.24 | -0.24 | -0.55; 0.06 | 0.12 |
|  | H-ST | -0.09 | -0.36; 0.17 | 0.48 | -0.18 | -0.50; 0.14 | 0.27 |
|  | N-CT | -0.15 | -0.42; 0.11 | 0.26 | -0.14 | -0.46; 0.18 | 0.37 |
|  | N-ST | - | - | - | - | - | - |
| Subjective cognition | H-CT | -0.10 | -0.37; 0.16 | 0.43 | -0.10 | -0.38; 0.18 | 0.50 |
|  | H-ST | -0.22 | -0.50; 0.05 | 0.12 | -0.13 | -0.43; 0.16 | 0.37 |
|  | N-CT | -0.09 | -0.36; 0.19 | 0.53 | -0.09 | -0.38; 0.20 | 0.54 |
|  | N-ST | - | - | - | - | - | - |
| Functioning | H-CT | -0.02 | -0.30; 0.26 | 0.88 | -0.21 | -0.52; 0.11 | 0.19 |
|  | H-ST | -0.25 | -0.55; 0.04 | 0.09 | -0.24 | -0.57; 0.09 | 0.15 |
|  | N-CT | -0.09 | -0.38; 0.21 | 0.57 | -0.05 | -0.38; 0.28 | 0.75 |
|  | N-ST | - | - | - | - | - | - |
| Sleep | H-CT | 0.15 | -0.20; 0.51 | 0.40 | -0.03 | -0.44; 0.38 | 0.88 |
|  | H-ST | 0.09 | -0.28; 0.46 | 0.63 | 0.23 | -0.21; 0.66 | 0.30 |
|  | N-CT | 0.03 | -0.35; 0.40 | 0.89 | -0.07 | -0.51; 0.36 | 0.74 |
|  | N-ST | - | - | - | - | - | - |
| **Note:** Outcomes are z-standardized based on the baseline means and standard deviations across the whole sample. Includes all available data (incl. study dropouts). <sup>1</sup>Missing data at baseline for N-CT: *n*=4 and N-ST: *n*=2; missing data at treatment completion for H-CT: *n*=1, N-CT: *n*=3, and N-ST: *n*=2; missing data at follow-up for N-CT: *n*=3 and N-ST: *n*=2. **Abbreviations:** CAVIR=Cognition Assessment in Virtual Reality; CI=Confidence interval; H-CT=Hypoxia with cognitive training; H-ST=Hypoxia with sham training; N-CT=Normoxia with cognitive training; N-ST=Normoxia with sham training. |  |  |  |  |  |  |  |

There was also a positive effect of N-CT (vs. N-ST) on ‘processing speed’, ‘verbal memory’, and ‘global cognition’ at treatment completion (*ps*≤0.03), of which improvements in verbal memory and global cognition prevailed at follow-up (*ps*≤0.049). None of these effects on tertiary outcomes rendered statistically significant after adjustment for multiple comparisons. Results for each individual tertiary outcome are reported in **Table S4**.

### Treatment fidelity and blinding

Participants completed *Mdn*=17 treatment sessions (IQR 16-18), with no difference across the four arms (*p*=0.33). Participants in the two cognitive training groups spent *M±SD*=30.9±5.8 hours on cognitive training, with no differences between groups (*p*=0.99), although participants who underwent cognitive training in normoxia vs. hypoxia generally achieved slightly higher difficulty levels across exercises (H-CT: *M±SD*=12.7±3.3; N-CT: *M±SD*=14.7±3.2, *p*=0.02, Cohen’s *d*=-0.64). For more details, see the **Supplement**.

After completion of the final follow-up assessment, 58% of participants in the H-CT group correctly guessed their allocated treatment, whereas it was 56% for the H-ST group, 71% for the N-CT group, and 54% of the N-ST group.

### Safety and tolerability

A total of four AEs were registered for three participants (H-ST: *n*=2; N-ST: *n*=1). These included moderate symptoms of headache, dizziness, and nausea. The two participants in hypoxia groups were excluded from the trial for safety reasons, while the participant in normoxia withdrew from the study. No serious AE was observed. As expected, hypoxia reduced mean SpO_2_ (hypoxia: *Mdn*=86.4% (IQR 84.7-87.6); normoxia: *Mdn*=97.5% (IQR 97.1-97.8), *p*<0.001, *d*=0.86) and increased pulse rates (hypoxia: *M±SD*=81.5±8.5, normoxia: *M±SD*=75.8±7.3, *p<*0.001, *d*=0.71). Haematological parameters were within reference values at all time points. There was a transient increase in haemoglobin and thrombocytes levels on days 8 and 19 for hypoxia vs. normoxia groups (*ps≤*0.04) that were normalized at follow-up (*ps*≥0.70) (**Table S8**; **Figure S5**). Reticulocytes levels were increased only on day 8 of hypoxia vs. normoxia (*p*=0.01), whereas erythrocytes levels were increased only on day 19 (*p*=0.02) with a numeric increase on day 8 (*p*=0.06). Participants reported none or very mild symptoms of altitude sickness on the ESQ, although hypoxia was generally associated with slightly higher total scores (hypoxia: *Mdn*=2.7 (IQR 1.6-5.2) (of possible 55), normoxia: *Mdn*=1.2 (IQR 0.5-3.1), *p*<0.001, *d*=0.31). Further details are provided in the **Supplement**.

### Mechanistic outcomes

#### Presynaptic density

There was a lower [^11^C]UCB-J *BP_ND_* in the hippocampus for H-CT vs. N-ST at treatment completion (H-CT: *M±SD*=2.58±0.39, N-ST: *M±SD*=2.85±0.22, *p*=0.03, *d*=-0.82) (**Figure 3A**), but no difference in the frontal cortex (*p*=0.22). Post hoc correlation analyses revealed a significant association between lower hippocampal *BP_ND_* and improvement in ‘global cognition’ from baseline to follow-up across both groups (*r*=-0.43, *p*=0.02; **Figure 3B**). There were no significant correlations between hippocampal *BP_ND_* and change in the other cognitive outcomes showing H-CT-related effects (*ps*≥0.10).

**Figure 3.**
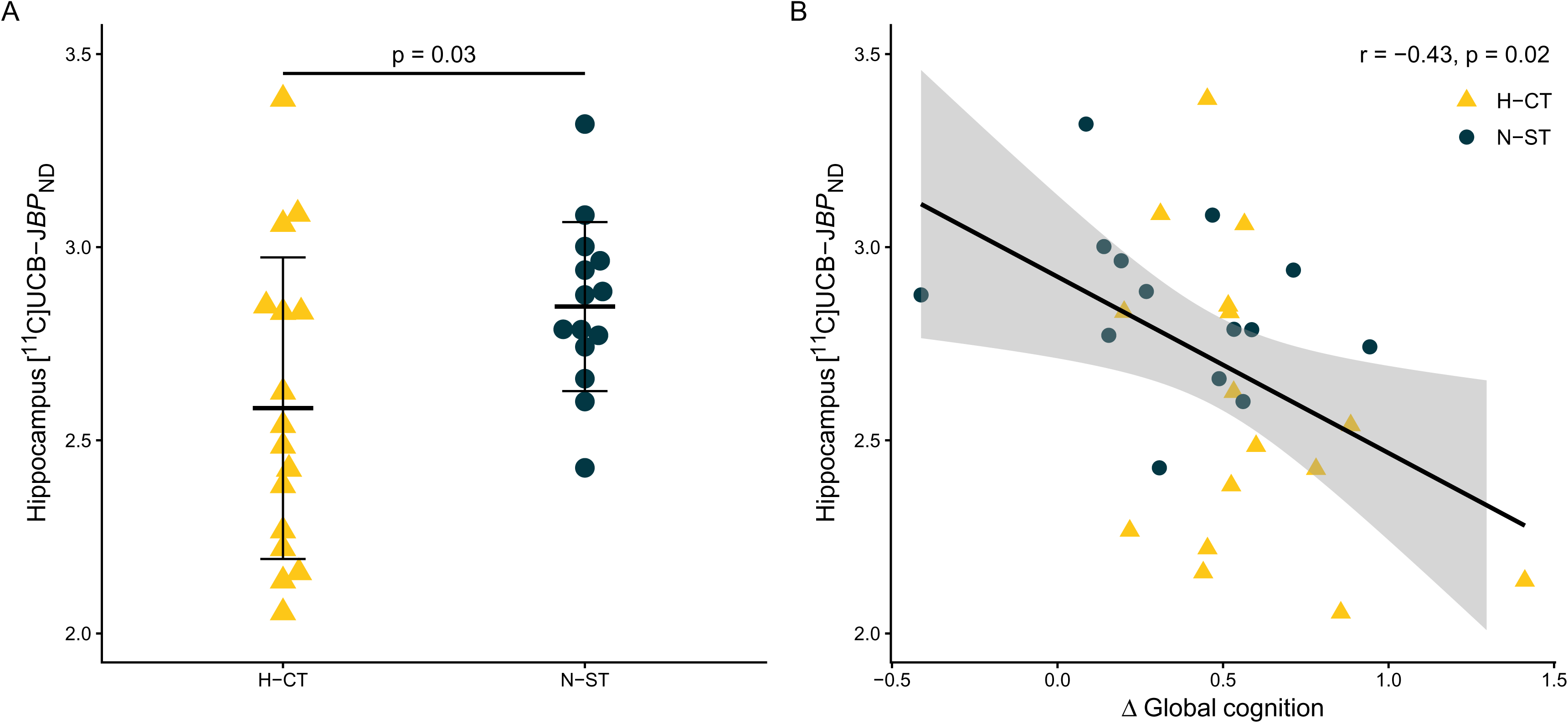
Effects of cognitive training under hypoxia on presynaptic density in the hippocampus at treatment completion (week 4). **Legend:** A: Comparison of synaptic vesicle glycoprotein 2A (SV2A) [^11^C]UCB-J binding in the hippocampus after three weeks of combined H-CT (*n*=16) versus sham training under normoxia (N-ST) (*n*=14). [^11^C]UCB-J non-displaceable binding potential (*BP_ND_*) was quantified using a simplified reference region model 2 (SRTM2) with centrum semiovale (white matter) as reference region. The crossbars show the means and standard deviations. B: Relationship between hippocampal *BP_ND_* and improvements in global cognition from baseline to one-month follow-up (week 8) across both groups (*n*=30). The shaded grey area represents the 95% confidence intervals. There was no significant association hippocampal *BP_ND_* and improvement in global cognition from baseline to treatment completion (week 4) (data not shown). Abbreviations: H-CT=Hypoxia with cognitive training; N-ST=Normoxia with sham training.

Group means and comparisons for the other exploratory regions are listed in **Table S9**. Of these, H-CT (vs. N-ST) also showed significantly lower *BP_ND_* in the following cortical regions: orbitofrontal gyrus, gyrus rectus, lingual gyrus, and cuneus (*ps*≤0.04). Post hoc correlation analyses showed associations between only lower *BP_ND_* in the gyrus rectus and improvements in ‘global cognition’ from baseline to follow-up (*r*=-0.52, *p*=0.003) and ‘attention’ from baseline to follow-up in both groups (*r*=-0.45, *p*=0.01).

#### Neural activity

Exploratory fMRI analyses showed no treatment effects on neural activity in the right DLPFC ROI during high-load working memory (*ps*≥0.37), nor were there any treatment-by-time interaction effects on working memory-related activity within the dorsal prefrontal cortex volume of interest or across the whole brain. Accordingly, there were no treatment effects on the in-scanner task performance (*d’* value) across the conditions (*ps*≥0.14).

#### Peripheral biomarkers

There was a non-significant numeric decrease in serum EPO from baseline to day 19 for the H-CT (*p*=0.09) and H-ST (*p*=0.08) groups, relative to N-ST (**Table S10**). There was a significant decrease in serum BDNF in the N-CT (vs. N-ST) group from baseline to day 19 (treatment effect=-0.62, 95% CI=[-1.20;-0.13], *p*=0.01), and a non-significant numeric decrease for the H-CT group (*p*=0.06). There was no treatment effect on serum VEGF (*ps*≥0.10).

When exploratorily combining hypoxia (vs. normoxia) groups, there was a significant decrease in EPO (treatment effect=-0.25, 95% CI=[-0.42;-0.78], *p*=0.01) and increase in VEGF (treatment effect=0.20, 95% CI=[0.01;0.39], *p*=0.04), but no change in BDNF (*p*=0.72) (**Table S11**). When exploratorily combining both cognitive training (vs. sham) groups, there was a decrease in BDNF (treatment effect=-0.42, 95% CI=[-0.75;-0.10], *p*=0.01), but no effects on EPO or VEGF (*ps*≥0.66). Post hoc analyses revealed no significant correlations between changes in EPO, VEGF, or BDNF and change in the cognitive outcomes that showed H-CT-related effects (*ps*≥0.06).

## Discussion

This is the first trial to investigate the combined and separate effects of moderate hypoxia (12% O₂) and cognitive training over three weeks on cognition and neuroplasticity in 126 healthy individuals. The primary cognitive composite outcome, ‘speed of complex cognitive processing’, was not improved by H-CT at treatment completion, and the trial must therefore be declared formally negative. However, H-CT showed efficacy on this cognitive outcome one month post-treatment, pointing to a potential delayed effect. As hypothesised, cognitive training alone (N-CT) yielded only immediate and transient effects on the primary outcome, and hypoxia without training (H-ST) elicited no significant change. Together, these overall results could suggest that a multimodal intervention combining physiological and cognitive stressors is essential for inducing long-term neurocognitive benefits.

Contrary to our primary hypothesis, the cognitive effects of H-CT had not yet appeared at treatment completion but emerged one month later. This delayed response may be attributable to several factors. Firstly, it cannot be excluded to be a chance finding. On the other hand, several other observations in the study suggest that the delayed response is a true finding. Newly matured neurons typically require four weeks for full functional integration in their neurocircuitries, which is essential for the cognitive benefits to emerge [50]. While neuroplastic alterations occurred during H-CT, as demonstrated by the lower hippocampal presynaptic density measured with [^11^C]UCB-J PET at treatment completion, any possible new neuronal connections were unlikely to have become functionally active at this time. Consistent with this interpretation, lower hippocampal [^11^C]UCB-J *BP_ND_* was associated with greater improvements in global cognition after one month, suggesting that these earlier synaptic changes may precede and facilitate subsequent cognitive gains. This pattern further aligns with a previous clinical study showing memory improvement alongside reversal of hippocampal volume loss *one month* after exogeneous EPO treatment [29]. Another explanation may be that the dual H-CT intervention imposed additional physiological strain because of the hypoxia, as evidenced by rises in red blood cell count, reduced training performance during hypoxia exposure, and increased self-reported fatigue immediately after sessions compared to normoxia. These mild stressors may have temporarily constrained acute cognitive gains and delayed their emergence. In line with this interpretation, the combined intervention could also have triggered a hormetic response, whereby mild stress combined with adequate physiological recovery post-intervention induced adaptive, compensatory neuroplasticity mechanisms that enhanced cognition over time [51]. This may also explain the absence of associations among change in peripheral biomarkers (EPO, VEGF, and BDNF) during treatment and subsequent cognitive improvements, since key mechanisms underlying hormetic effects likely emerge *after* the treatment period, though it is also possible that these biomarkers were not directly related to the observed cognitive benefits.

Conversely, we observed no treatment-related change in our co-secondary outcome, working memory-related DLPFC activity, one month post-treatment. This contrasts with prior studies demonstrating DLPFC engagement in response to different pro-cognitive interventions in psychiatric populations (e.g., [27, 38]). One possible explanation is that those studies reported improvements in executive or verbal memory functions, which are closely linked to DLPFC integrity. In contrast, H-CT elicited no change in executive functions (co-secondary outcome) but predominantly enhanced broad cognitive composite measures and attention specifically (primary and tertiary outcomes, respectively). These aspects of cognition rely on more distributed neural networks, which are not directly captured by the working memory N-back task. It is therefore possible that H-CT preferentially targets neurocircuitries involved in these distinct cognitive processes, a hypothesis that requires further investigation.

The direction of observed difference in presynaptic density for H-CT versus N-ST groups was surprising, since previous [^11^C]UCB-J PET studies have shown a link between lower SV2A levels and *poorer* cognitive performance in psychiatric samples [52–54]. Our findings could suggest that this relationship potentially differs in the healthy brain, since lower [^11^C]UCB-J *BP_ND_* was associated with greater cognitive benefits. Specifically, H-CT might trigger adaptive synaptic pruning processes, whereby superfluous or ‘less-active’ synapses are eliminated, and new neurons are integrated into existing, refined circuits [55, 56], resulting in greater long-term cognitive improvement. Other mechanisms may involve the increase in VEGF observed across H-CT and H-ST groups, which aligns with previous findings that HIF-stabilization enhances VEGF signalling [16, 18, 57] - this pathway has been causally linked to increased neurogenesis in rodents [57]. Similarly, both exogeneous and endogenous EPO have previously demonstrated neuroprotective effects [10, 29, 58], and the observed serum EPO reduction over time in hypoxia-treated participants, compared to those in normoxia, was therefore unexpected. This decrease may reflect a hormonal feedback mechanism: initial hypoxia-induced EPO stimulates erythropoiesis – as evidenced by the early rise in red blood cell count after one week of treatment – which in turn triggers negative feedback, suppressing further EPO production and leading to its reduction by day 19 [59]. This pattern is consistent with a single-arm pilot study using a similar hypoxia protocol showing that serum EPO peaked after one week and slightly declined thereafter [60]. The decrease in serum BDNF after cognitive training may also reflect similar feedback mechanisms, particularly considering the observed benefits of the training. However, our finding contrasts with previous reports of increased BDNF after cognitive training (e.g., [61, 62]). To better elucidate these dynamics, future studies should incorporate early and more frequent sampling during the intervention.

Strengths include the randomised, controlled, four-arm design with well-matched groups. We further adopted a multidisciplinary translational approach to elucidate the mechanisms underlying changes in neuroplasticity and cognition across multiple levels of analysis, including presynaptic density, peripheral biomarkers, neurocircuitry activity, and cognitive tests. A possible limitation is that the relatively small treatment effects on cognition may be attributable to near-ceiling effects in our sample, as there is less room for improvement in cognitively intact, young healthy individuals. We did not assess possible effects of H-CT beyond one month after treatment completion, so longer-term effects must be investigated in future studies. Double-blinding was successful in three of the four groups but less effective in the N-CT group, potentially introducing expectancy effects that may have influenced cognitive outcomes. Finally, our hypoxia protocol involved externally adjusting the fraction of inspired oxygen (FiO_2_) to 12%, based on our prior preclinical work [10] and for feasibility reasons. However, future studies should consider providing individually adjusted hypoxia to achieve a comparable SpO2 across participants to reduce interindividual variability [63,64].

In conclusion, while there was no significant effect on the primary endpoint, our study suggests that three weeks of cognitive training under moderate hypoxia produces delayed cognitive improvements in healthy humans after one month. H-CT was accompanied by reduced hippocampal presynaptic density, which correlated with long-term cognitive gains. In contrast, normoxic cognitive training produced only a transient cognitive improvement that disappeared after one month. Combining hypoxia with cognitive training may represent a promising strategy for enhancing cognitive proficiency, likely through modulation of hippocampal synaptic plasticity and neuronal transcription factors. Future research should investigate the clinical utility of H-CT in neuropsychiatric populations, for whom conventional interventions yield limited cognitive benefits.

## Supporting information

CONSORT checklist

## Data Availability

Deidentified data collected and presented in this study is available from the corresponding author (KWM) upon reasonable request.

## Acknowledgements

We would like to thank all participants for dedicating their time and effort to take part in the study. We also thank medical students, Barbara Emilie Rotbøl Ørum and Anne Bügel Fisker Madsen, and the research assistants in the NEAD Centre for their valuable work with neuropsychological testing, treatment monitoring, and fMRI scanning. The technical assistance of Nadine Becker von-Buch is also gratefully acknowledged. The study was funded by the European Research Council (ERC) Consolidator Grant awarded to PI Kamilla Miskowiak (grant no. 101043416). In addition, the study has received financial support from the AP Møller foundation (grant no. 2022-00134). Medical students, Ida Plougmann Østergaard and Katrine Krabbe Thommesen, received financial support from the Lundbeck Foundation (grant no. R464-2024-122 and R405-2022-87, respectively) to work on the project during their research internship. The study funders had no role in the study design, data collection and analysis, and writing of the article.

## Author contributions statement

Conceptualization: KWM, HE. Data curation: VD, JMS, KC, IPØ, KKT, CFB, MBJ, LVK. Methodology: VD, JMS, AJ, JM, PPS, SL, CS, GMK, MBJ, LVK, HE, KWM. Formal analysis: VD, JMS, AJ, JM, MM, CS, KWM. Writing – original draft: VD, KWM. Writing – review and editing: All authors.

