## Supplement for "Effects of cognitive training under inspiratory hypoxia on cognition and neuroplasticity in healthy humans: a randomised, double-blind, controlled, four-arm trial"

### SUPPLEMENT

### METHODS

#### Trial preregistration

The present trial in healthy humans and a parallel study in people with mood disorders were jointly preregistered together on ClinicalTrials.gov (NCT06121206) on the 31st October 2023. Registration was completed retrospectively due to delays in obtaining approval from the local ethics committee, which arose from amendments to the design in the mood disorder study only. However, registration was prospective for all outcomes in the present study involving healthy participants.

#### Power calculation

The power calculation was based on the primary hypothesis that combined H-CT vs. N-ST produces cognitive improvement (hypothesis i). As hypoxia triggers EPO production, the power calculation was based on our prior clinical study of exogenous EPO treatment that showed a differential change of 0.5 SD between EPO and placebo groups in the same cognitive composite score as we apply here [1]. We assumed an effect of 0.4 *z*-scores (medium effect size) differential change between the two groups with 0.5 SD of the treatment effect across groups. To achieve >80% power to identify this effect on the primary outcome at an α-level of 0.05, a total of 104 participants (26 per arm) were required. To accommodate an expected 20% attrition rate, we included 30 participants per group (120 in total). After inclusion of the 120 participants, we enrolled an extra six (also blinded) participants allocated to the H-CT group to obtain a sufficient sample size for the PET imaging analysis (see details below).

#### Eligibility criteria

Inclusion criteria were age between 18 and 50 years and fluent in Danish. Exclusion criteria were current or prior psychiatric illness confirmed using the Schedules for Clinical Assessment in Neuropsychiatry (SCAN) interview [2], neurological illness or history of serious head trauma, alcohol or substance abuse, and dyslexia. To ensure safety, participants were also excluded if they had prior experience of altitude sickness, current or prior somatic conditions (i.e., heart disease, lung disease, kidney disease, diabetes, untreated or insufficiently treated hypertension, and thrombosis or first-degree family with thromboembolic events prior to age 60), pregnancy or breastfeeding, smoking or other regular use of nicotine products, current use of iron supplementation, and body mass index (BMI)>30. Participants were not eligible for PET if they had participated in experiments with radioactivity (>10 mSv) over the past year, had significant occupational exposure to radioactivity, or took incompatible medication (i.e., synaptic vesicle glycoprotein 2A (SV2A) binding agents). For MRI, participants were further excluded if they suffered from claustrophobia, had a pacemaker, or had other contraindications for MRI. Participants were comprehensively screened to ensure compliance with inclusion and exclusion criteria, involving a semi-structured interview, questionnaires, blood sampling, electrocardiogram (ECG), measurement of blood pressure, lung and heart auscultation, and assessment of weight and height. Participants were not screened for prior or habitual exposure to hypoxic environments; however, it is considered unlikely that any participant was acclimatized to hypoxia before beginning the intervention given Denmark's low altitude.

**Treatment sessions**

The treatment sessions were conducted at the Department of Psychology, University of Copenhagen from February 2023 to May 2025. The treatment involved 3.5-hour sessions, six days per week (Mondays-Saturdays) for three weeks (18 sessions in total). In cases of sickness unrelated to the treatment, participants were offered a booster session immediately in extension of the three-week intervention. During the treatment sessions, 2-4 participants stayed in the altitude training room where they each sat at a desk separated by desk dividers (**Figure S1** shows the experimental setup). All participants wore a pulse oximeter that continuously monitored their $SpO_2$ and pulse rate (Shanghai Berry Electronic Tech Co., Ltd). The first 30 minutes was a lead-in phase, where the oxygen levels were either lowered from 16% to 12% for the hypoxia groups or were stable at 20% for the normoxia groups, and where the participants relaxed. They then performed cognitive training or matched sham games on an iPad for two hours each treatment session. The cognitive or sham training was interleaved with short breaks inside the altitude room, where the participants relaxed (3 x 10 minutes) or slowly walked on an in-room treadmill (max 3 km/hour) (3 x 10 minutes) to counteract possible negative effects of sedentary behaviour on cognition [3]. In the middle of the treatment session, participants had a short bathroom break, where they leaved the altitude room (approx. 2 minutes). Water, coffee, tea, and snacks were available for the participants during the treatment sessions. Caffeine intake during treatment sessions was permitted to better reflect real-world conditions and was therefore not recorded. However, participants were instructed to refrain from consuming caffeine prior to all assessments. Before and after each treatment session, participants were interviewed about their sleep and physical well-being. Participants also completed a Visual Analogue Scale (VAS) questionnaire about their subjective state and any adverse reactions before and after each treatment session, in addition to cerebral symptoms of altitude sickness with the Environmental Symptoms

Questionnaire [4] (ESQ) after each session. During the treatment sessions, unblinded research personnel monitored participants' $SpO_2$, pulse rate, and subjective states from the questionnaires.

#### Cognitive and sham training

The cognitive training was provided by the web-based program, Happy Neuron Pro (www.research.scientificbraintrainingpro.eu, Danish version, 2024). The program consisted of 26 different exercises that tap into wide-spread cognitive domains, including auditory processing, processing speed, attention, working memory, visual and verbal memory, and executive functions. The exercises were designed to be engaging, motivating, and relevant to everyday functioning. For the active training condition, each exercise had 30 difficulty levels that increased following consecutive trials with 80% accuracy. The active training further involved parametric task adjustment by decreasing stimuli presentation time, increasing working memory load, decreasing time to respond, and increasing the number of non-target items. For the sham training condition, participants completed similar exercises but with low cognitive demand. The sham training included the exact same stimuli as the cognitive training condition, but with no adjustments from trial to trial except in the appearance of the exercises. All participants were instructed to train for two hours per session.

**Figure S1:** Experimental setup of the altitude training room and cognitive training conducted at the Department of Psychology, University of Copenhagen. Each treatment session was 3.5 hours and included iPad-based cognitive training interleaved with breaks where participants either relaxed or walked slowly on a treadmill. Participants wore a pulse oximeter during treatment sessions (display not visible to participants to maintain blinding). The current $O_2$ level and each participant's $SpO_2$ and pulse rate were continuously monitored by unblinded research personnel outside the altitude training room.

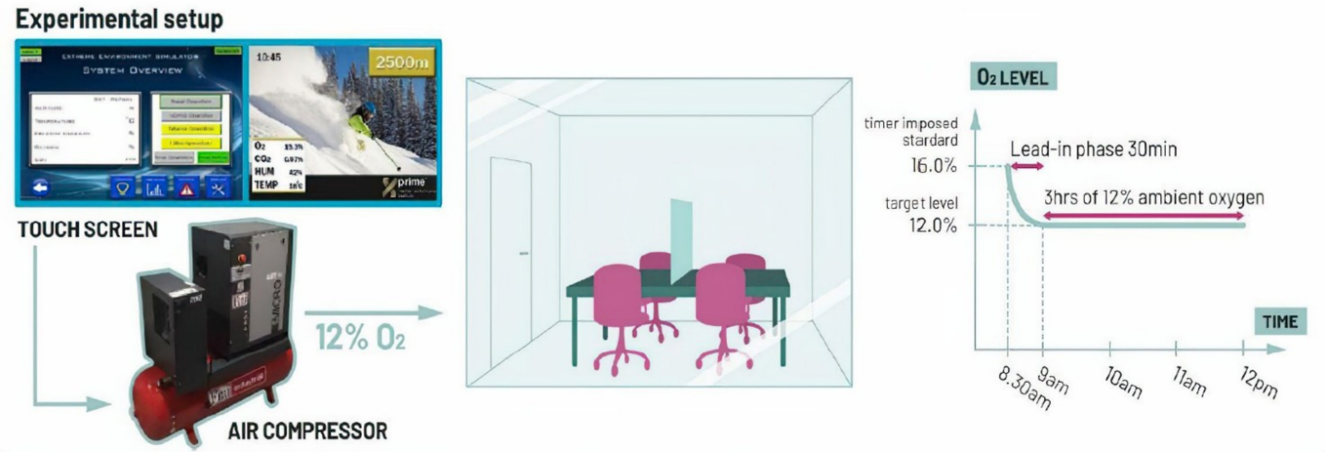

**Figure S2**: Overview of study procedures. Participants underwent screening for inclusion and exclusion criteria prior to enrolling in the trial. Baseline assessments of cognition, magnetic resonance imaging, and routine blood samples were conducted the week prior to beginning the intervention. Additional blood samples (routine and biomarker) were collected during the three-week intervention. Participants were assessed again at treatment completion (week 4), where a subsample of participants also underwent positron emission tomography, and one month post-treatment (week 8). Abbreviations: BDNF = Brain-derived neurotrophic factor; EPO = Erythropoietin; fMRI = Functional magnetic resonance imaging; MRI = Magnetic resonance imaging; PET = Positron emission tomography; VEGF = Vascular endothelial growth factor.

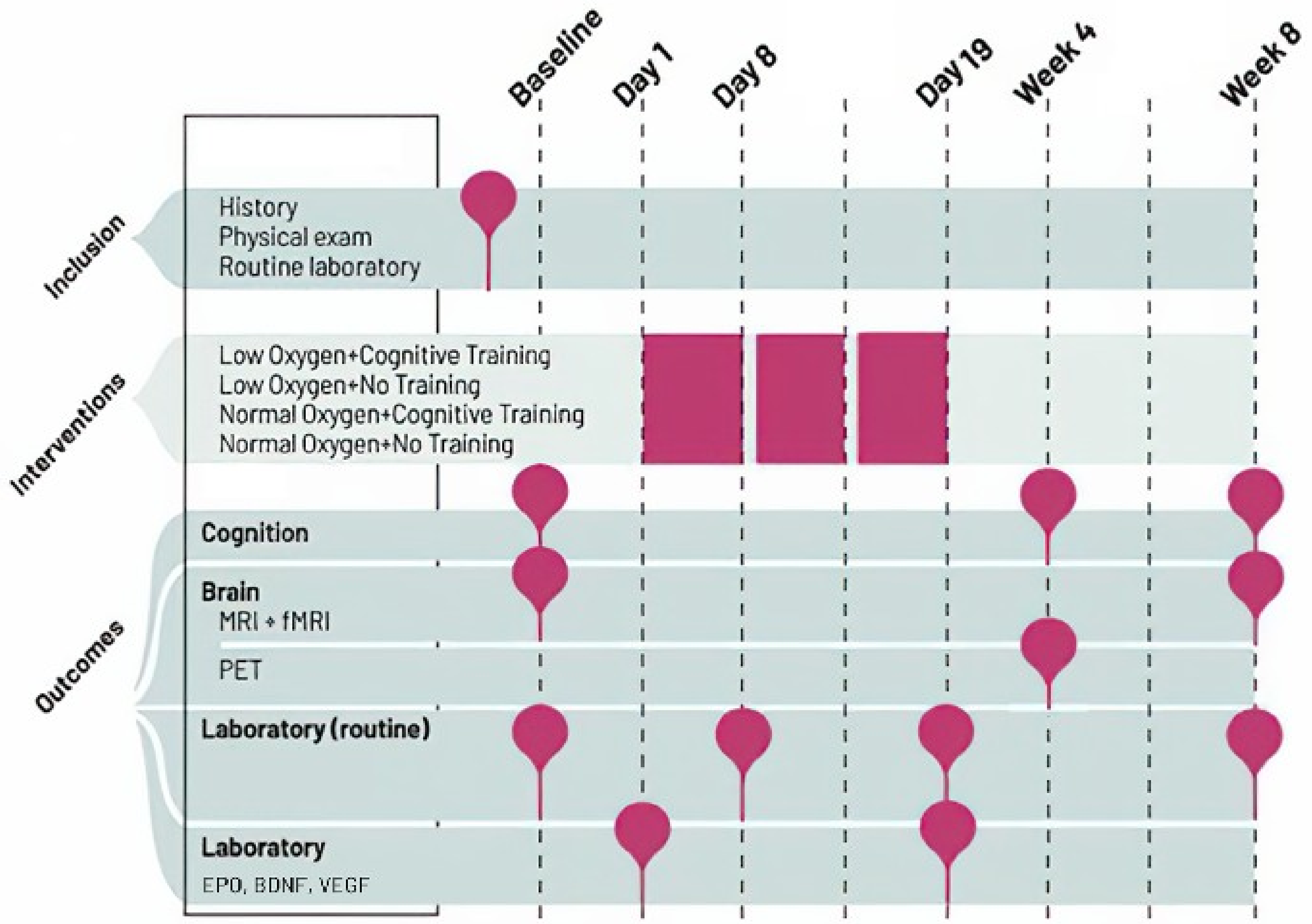

#### Outcome measures

For an overview of each primary, secondary, tertiary, and mechanistic outcome and their associated hypotheses, see **Table S3**.

##### *Primary outcome*

The primary outcome was a cognitive composite score, 'Speed of complex cognitive processing', based on an average of *z*-transformed scores from the following tests: Rey Auditory Verbal Learning Test (RAVLT) lists I-V total recall, Repeatable Battery for the Assessment of Neuropsychological Status (RBANS) Coding, Verbal Fluency (letter "D"), Wechsler Adult Intelligence Scale (WAIS)-III Letter-Number Sequencing, Trail Making Test Part B, and Rapid Visual Information Processing (RVP) from CANTAB (Cambridge Cognition Ltd.). To minimize learning effects from repeated testing, matched alternate versions of the RAVLT and RBANS Coding were used in a counter-balanced order. We selected this composite score as our primary outcome based on our previous study showing improvements in this measure after exogeneous erythropoietin (EPO) treatment [1]. The predefined endpoint was change in 'speed of complex cognitive processing' from baseline to treatment completion (week 4), whereas change from baseline to one-month follow-up (week 8) was assessed more exploratorily.

##### *Secondary outcomes*

The co-secondary cognitive outcome was 'Mean choices to correct' from One Touch Stockings of Cambridge (OTS; CANTAB). This is a measure of planning abilities (executive functions). We selected this co-secondary outcome based on our previous trial showing selective improvements in this score after similar cognitive training [5], which was also accompanied by neuroplastic changes during task functional magnetic resonance imaging (fMRI) [6]. The predefined endpoint was change in 'Mean choices to correct' from baseline to treatment completion (week 4), whereas change from baseline to one-month follow-up (week 8) was assessed more exploratorily.

The co-secondary neural outcome was 'dorsal prefrontal cortex response' during working memory fMRI. Specifically, we assessed change in the right dorsolateral prefrontal cortex (DLPFC) during a spatial working memory N-back task. A right DLPFC region of interest (ROI) was constructed eight mm around peak Montreal Neurological Institute (MNI) coordinates x=40, y=34, z=29, based on prior studies showing changes in this region after various pro-cognitive interventions, including exogeneous EPO treatment and cognitive training [6–8]. Specifically, the right DLPFC was chosen due to the paradigm's visuospatial component, primarily tapping into mainly the right hemisphere. We selected this co-secondary neural outcome, as we hypothesized that treatment-related cognitive improvements at the behavioural level would be accompanied by neuroplasticity changes captured at the neural level, possibly reflecting a biomarker for pro-cognitive effects [9]. Specifically, the predefined endpoint was change in mean percent blood-oxygen-level-dependent (BOLD) signal change in the right DLPFC ROI from baseline to follow-up (week 8) during the general working memory (2-back minus 0-back contrast).

***Tertiary outcomes***

Tertiary cognitive outcomes included change in global and domain-specific performance based on the full neuropsychological test battery: Rey Auditory Verbal Learning Test (RAVLT), Repeatable Battery for the Assessment of Neuropsychological Status (RBANS) Coding and Digit Span, Verbal Fluency with the letters 'S' and 'D', Wechsler Adult Intelligence Scale (WAIS)-III Letter-Number Sequencing, Trail Making Test Part A and B, the Wisconsin Card Sorting Task (WCST), additional measures from the One Touch Stockings of Cambridge (CANTAB), Rapid Visual Processing (CANTAB), Spatial Working Memory (SWM; CANTAB), Emotion Recognition Test (ERT; CANTAB), and the Cognition Assessment in Virtual Reality (CAVIR) test, an immersive VR tests probing different cognitive functions across five subtasks: 1) verbal memory, 2) executive functions, 3) processing speed, 4) working memory, and 5) attention [10]. To reduce possible learning effects, matched alternate versions of the RAVLT, RBANS Coding, and CAVIR were used in a counter-balanced order. Tertiary self-reported outcomes were total scores on the World Health Organization Quality of Life (WHOQOL-BREF), the Assessment of Quality of Life (AQoL), the Cognitive Complaints in Bipolar Disorder Rating Assessment (COBRA), the Pittsburgh Sleep Quality Index (PSQI), the Work and Social Adjustment Scale (WSAS), and Sheehan Disability Scale (SDS). For full list of all tests included in each domain, see **Table S2**.

**Table S2:** Overview of cognitive and self-report domains and each measure included.

| **Cognitive domain** | **Neuropsychological test** |
|---|---|
| Processing speed | RBANS Coding, Trail Making Test A |
| Attention | RBANS Digit Span, CANTAB RVP 'accuracy', 'mean latency', 'false alarms', and 'probability of hit' |
| Working memory | WAIS-III Letter-number sequencing, CANTAB SWM 'between errors' and 'strategy' |
| Verbal learning and memory | RAVLT total recall on subtests 'list I-V', 'immediate recall', and 'delayed recall' |
| Executive functions | Trail Making Test B, Verbal fluency total letters 'S' and 'D', CANTAB OTS 'problems solved on first choice' and 'mean latency to correct', Wisconsin Card Sorting Task 'perseverative errors' |
| Global cognition | Average score of the following domains: processing speed, attention, working memory, verbal learning and memory, and executive functions. |
| Facial expression recognition | CANTAB ERT unbiased 'hit rate' (average across all six emotions) and 'median reaction time' across all trials |
| CAVIR global composite | A combined measure of CAVIR subtasks 1-5 as described in [10]. |
| **Self-report domain** | **Questionnaire** |
| Quality of life | World Health Organization Quality of Life (WHOQoL-BREF), Assessment of Quality of Life (AQoL) |
| Subjective cognition | Cognitive Complaints in Bipolar Disorder Rating Assessment (COBRA) |
| Functioning | Work and Social Adjustment Scale (WSAS), Sheehan Disability Scale (SDS) |
| Sleep | Pittsburgh Sleep Quality Index (PSQI) |

**Abbreviations:** CAVIR=Cognition Assessment in Virtual Reality; ERT=Emotion Recognition Test; OTS=One Touch Stockings of Cambridge; RAVLT=Rey Auditory Verbal Learning Test; RBANS= Repeatable Battery for the Assessment of Neuropsychological Status; RVP=Rapid Visual Information Processing Test; SWM=Spatial Working Memory; WAIS=Wechsler Adult Intelligence Scale.

***Mechanistic outcomes***

Mechanistic PET outcomes were difference in synaptic vesicle glycoprotein 2A (SV2A) [$^{11}$C]UCB-J binding as a readout of presynaptic density between H-CT and N-ST groups at treatment completion (week 4). Although the PET outcomes were mechanistic and thus conducted for exploratory purposes, our a priori measures of interest were non-displaceable binding potential ($BP_{\mathrm{ND}}$) in the bilateral hippocampus (key hub for neurogenesis) and bilateral frontal cortex (involved in general cognitive proficiency). To explore any unpredicted change in presynaptic density outside these regions, we also estimated $BP_{\mathrm{ND}}$ in the following 23 cortical ROIs: neocortex, insula, anterior cingulate cortex, posterior cingulate cortex, middle frontal gyrus, precentral gyrus, postcentral gyrus, gyrus rectus, orbitofrontal gyri, inferior frontal gyrus, superior frontal gyrus, anterior temporal lobe (medial part), anterior temporal lobe (lateral part), posterior temporal lobe, parahippocampal and ambient gyri, superior temporal gyrus, middle and inferior temporal gyri, fusiform gyrus, superior parietal gyrus, inferolateral remainder of parietal lobe, lingual gyrus, cuneus, lateral remainder of occipital lobe, and 6 subcortical ROIs: amygdala, nucleus accumbens, caudate nucleus, putamen, thalamus, and striatum. All ROIs were bilateral. The neocortex was defined as the weighted average of the individual subregions: frontal, parietal, temporal, occipital, and insular cortices.

Mechanistic fMRI outcomes included change in mean percent BOLD signal change in the right DLPFC ROI during high-load specific working memory (i.e., 2- minus 1-back contrast) from baseline to one-month follow-up, in addition to treatment-related change over time across the whole dorsal prefrontal cortex and whole brain during both contrasts.

To further aid insights into the underlying neurobiological mechanisms involved in treatment-related cognitive changes, we exploratorily analysed blood samples collected at baseline and second to last day of treatment (day 19) for peripheral biomarkers, including serum concentrations of EPO, vascular endothelial growth factor (VEGF), and brain-derived neurotrophic factor (BDNF). The specific purpose was to investigate treatment-related change from baseline to day 19 in these biomarkers and then assessing if change in these markers correlate with change in cognition.

**Table S3**: Overview of outcomes and associated hypotheses.

| Outcome | Hypothesis | Timepoints |
|---|---|---|
| **Primary** | | |
| 'Speed of complex cognitive processing' composite measure | **(1)** H-CT (vs. N-ST) produces immediate improvement in 'speed of complex cognitive processing'<br>**(2)** ... that sustains one month post-treatment.<br><br>**(3)** H-ST and N-CT (vs. N-ST) produce immediate improvement in 'speed of complex cognitive processing' that does not sustain one month post-treatment. | **(1)** BSL-W4 (pre-defined primary outcome endpoint)<br>**(2)** BSL-W8 (exploratory primary outcome endpoint)<br><br>**(3)** BSL-W4+BSL-W8 (exploratory) |
| **Secondary** | | |
| OTS 'Mean choices to correct' | **(1)** H-CT (vs. N-ST) produces immediate improvement in executive functions, mimicking the selective positive effects of cognitive training observed in our prior trial [5]<br>**(2)** ... that sustains one month post-treatment. | **(1)** BSL-W4 (pre-defined secondary outcome endpoint)<br>**(2)** BSL-W8 (exploratory secondary outcome endpoint) |

| | | |
|---|---|---|
| | **(3)** H-ST and N-CT (vs. N-ST) produce immediate improvement in executive functions that does not sustain one month post-treatment. | **(3)** BSL-W4+BSL-W8 (exploratory) |
| DLPFC activation during working memory fMRI | **(1)** H-CT (vs. N-ST) increases task-related DLPFC activation one month post-treatment, in line with the neural effects of exogeneous EPO observed in our prior clinical trials [8, 11].<br><br>**(2)** H-ST and N-CT (vs. N-ST) do not increase task-related DLPFC activation one month post-treatment. | **(1)** BSL-W8 (pre-defined secondary outcome endpoint)<br><br>**(2)** BSL-W8 (exploratory) |
| **Tertiary** | | |
| Global and domain-specific cognitive composite scores | **(1)** H-CT (vs. N-ST) produces immediate and sustained cognitive improvement.<br><br>**(2)** H-ST and N-CT (vs. N-ST) produce immediate cognitive improvement that does not sustain one month post-treatment. | **(1)** BSL-W4+BSL-W8 (pre-defined tertiary outcome endpoints)<br><br>**(2)** BSL-W4+BSL-W8 (exploratory) |
| Self-report composite scores | **(1)** H-CT (vs. N-ST) produces immediate and sustained self-reported improvement in quality of life, subjective cognition, functioning, and sleep.<br><br>**(2)** H-ST and N-CT (vs. N-ST) produce immediate, but not sustained, self-reported improvement in quality of life, subjective cognition, functioning, and sleep. | **(1)** BSL-W4+BSL-W8 (pre-defined tertiary outcome endpoints)<br><br>**(2)** BSL-W4+BSL-W8 (exploratory) |
| **Mechanistic** | | |
| [$^{11}$C]UCB-J $BP_{ND}$ | **(1)** H-CT (vs. N-ST) produces immediate change in presynaptic density in the hippocampus and frontal cortex, similar to the neuroplastic effects of exogeneous EPO and/or hypoxia seen in [12, 13]. | **(1)** BSL-W4 |
| Dorsal prefrontal cortex activation + DLPFC activation during high-load specific working memory fMRI | **(1)** H-CT (vs. N-ST) increases task-related dorsal prefrontal cortex activation one month post-treatment.<br><br>**(2)** H-ST and N-CT (vs. N-ST) do not increase task-related dorsal prefrontal cortex activation one month post-treatment. | **(1)** BSL-W8<br><br>**(1)** BSL-W8 |
| Serum EPO, VEGF, and BDNF | **(1)** H-CT (vs. N-ST) increases serum EPO, VEGF, and BDNF.<br><br>**(2)** H-ST increases serum EPO and VEGF, whereas N-CT increases only serum BDNF. | **(1)** BSL-D19<br><br>**(2)** BSL-D19 |

**Abbreviations:** BDNF=Brain-derived neurotrophic factor; BPND=Non-displaceable binding potential; BSL=Baseline; D19=Day 19; DLPFC=Dorsolateral prefrontal cortex; EPO=Erythropoietin; fMRI=Functional magnetic resonance imaging; H-CT=Hypoxia with cognitive training; H-ST=Hypoxia with sham training; N-CT=Normoxia with cognitive training; N-ST=Normoxia with sham training; OTS=One Touch Stockings of Cambridge; VEGF=Vascular endothelial growth factor; W4=Week 4; W8=Week 8.

##### *Statistical analysis of outcome measures*

Raw cognitive test or self-report scores were *z*-standardized based on the baseline means and standard deviations across the whole sample. Domain scores were computed by averaging *z*-scores. A 'global cognition' score was calculated by averaging all cognitive domain scores.

To assess treatment effects on each outcome, we performed linear mixed effects models with an unstructured covariance pattern. The unstructured covariance pattern was selected to avoid assuming equal variance across time points – an assumption that we particularly expected not to hold at follow-up. This statistical approach is consistent with our previous trials [5] and was recommended by a consulting biostatistician. Fixed effects were time (baseline, treatment completion, and one-month follow-up), treatment (H-CT vs. H-ST vs. N-CT vs. N-ST), and their interaction (time*treatment). N-ST was entered as the reference group, and baseline constraint was applied. The constraint involved assigning all participants to the reference group (N-ST) at baseline.

To control the false discovery rate (FDR) for multiple comparisons among secondary and tertiary outcomes, the Benjamini-Hochberg procedure was applied. The FDR was set to 5%. *p*-values were ranked from smallest to largest and compared to their critical values, calculated as $(i/m)\times Q$, where $i$ is the rank, $m$ the total number of tests, and $Q$ the chosen FDR (i.e., 0.05). *p*-values smaller than or equal to their corresponding critical values were considered statistically significant. Adjusted *p*-values were also calculated, and results with adjusted $p<.05$ were considered statistically significant.

For predefined outcomes showing a significant effect of H-CT vs. N-ST, we exploratorily tested for possible synergistic effects of the combined interventions by conducting separate linear mixed models. Fixed effects were time (baseline, treatment completion, and one-month follow-up), hypoxia treatment (yes/no), cognitive training treatment (yes/no), and their interactions (time*hypoxia*cognitive training).

#### Positron emission tomography analysis

##### *Power calculation*

The power calculation for the PET imaging of SV2A [$^{11}$C]UCB-J binding as a readout of presynaptic density was based on the following two assumptions: (1) the average $BP_{\mathrm{ND}}$ in the frontal cortex in healthy humans is $M \pm SD = 3.36 \pm 0.38$, based on in-house data, and (2) combined H-CT will lead to a 10% increase in $BP_{\mathrm{ND}}$ in frontal cortex and/or hippocampus. To achieve >80% power to identify this group difference in $BP_{\mathrm{ND}}$ using a two-tailed, independent samples *t*-test with an α-level of 0.05, we aimed to include a total of 40 participants (20 per treatment group). However, due to technical issues, the PET scanner was temporarily unavailable during the trial period, and we therefore achieved a sample of 30 participants for statistical analysis (see below).

##### *PET subsample and missing data*

See **Figure S3** for flow diagram for the subsample included in the PET imaging analysis. Among the 66 participants allocated to the two extreme groups (H-CT and N-ST), 60 participants completed the treatment and post-treatment assessments. Of these, 30 participants (H-CT: $n$=17; N-ST: $n$=13) were not included in the PET analysis, either because they were included in the study prior to starting PET scans ($n$=4), did not want to or did not have time to undergo PET ($n$=7), the scanner was unavailable due to temporary closure ($n$=14), were not eligible for PET or MRI ($n$=2), there were issues with the tracer from the radiochemistry production ($n$=2), or there were technical issues during PET acquisition ($n$=1). This yielded a final subsample of 30 participants (H-CT: $n$=16; N-ST: $n$=14) for PET analysis.

Baseline demographic characteristics were well-balanced between the treatment groups for the subsample included in the PET imaging analysis, with no significant differences in age (H-CT: $M \pm SD = 28.1 \pm 7.6$, N-ST: $M \pm SD = 26.4 \pm 5.3$, $p$=0.48), sex (H-CT: 50% female, N-ST: 50% female, $p$>0.99), verbal IQ (H-CT: $M \pm SD = 109.4 \pm 6.6$, N-ST: $M \pm SD = 106.3 \pm 5.2$, $p$=0.16), and years of education (H-CT: $M \pm SD = 15.8 \pm 3.5$, N-ST: $M \pm SD = 14.5 \pm 2.9$, $p$=0.29). Finally, the PET subsample ($n$=30) did not significantly differ from the remaining participants that were not included in the PET analysis ($n$=96) on age, sex, IQ, and years of education ($ps \geq 0.42$).

**Figure S3:** CONSORT flow diagram with exclusion reasons for the subsample of participants in the H-CT (*n*=16) and N-ST (*n*=14) groups included in the PET imaging analysis.

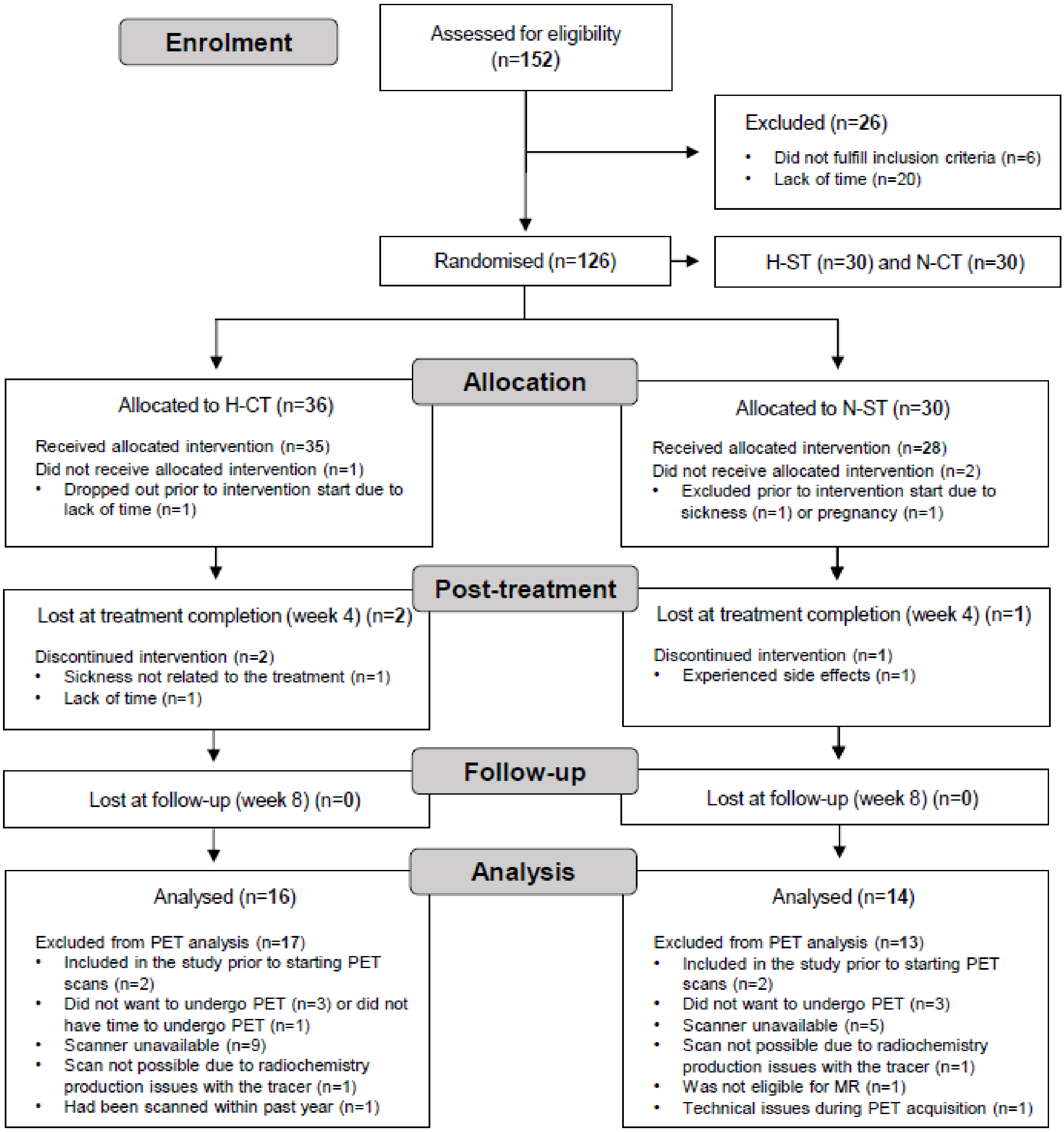

##### *Data acquisition*

Data were acquired the Copenhagen University Hospital, Rigshospitalet, Denmark using a high-resolution research tomography (HRRT) PET scanner (CTI/Siemens, Knoxville, TN, USA). Participants underwent a six-minutes transmission scan followed by a 90-minute emission scan, which started at the time of the intravenous bolus injection of between 100 and 510 MBq [$^{11}$C]UCB-J administered over 20 seconds. PET data were acquired in 3D list mode and reconstructed into 37 frames (8x15 s, 8x30 s, 4x60 s, 5x2 min, 10x5 min, and 2x10 min) using a 3D OP-OSEM algorithm with modelling of the point-spread-function, and attenuation corrected using the HRRT maximum a posteriori transmission reconstruction method (MAP-TR). Each image frame consisted of 207 planes of 256 × 256 voxels of 1.22 × 1.22 × 1.22 $mm^3$.

##### *Pre-processing*

All PET images were motion corrected using the Automated Image Registration (AIR) software with the reconcile command (v. 5.2.5) [14]. The remaining pre-processing was conducted using the PVElab pipeline (Neurobiology Research Unit, Copenhagen; https://nru.dk/pveout/). Specifically, PVElab used an unfiltered summation PET image that was automatically co-registered to the same participant's corresponding follow-up T1-weighted MR image using SPM12. Multispectral segmentation (i.e., using both T1 and T2-weighted MR images) was then used to extract time-activity curves from each automatically defined ROI [15–17]. Accurate co-registration and ROI placement was confirmed by visual inspection across all planes. No manual correction was needed. No partial volume effects correction was applied. The centrum semiovale (white matter reference region) was obtained from the PVElab region and was further eroded twice with a 3D erosion operator to minimize partial volume effects. The final volume was $M \pm SD$=7.45±2.63 mL.

##### *Kinetic modelling*

Regional time-activity curves were fitted to the simplified reference tissue model 2 (SRTM2) to estimate $BP_{\text{ND}}$ using the white matter region, centrum semiovale, as a reference region [18, 19]. For each fit, we used a population based k2' value of 0.035 min−1, based on the median value estimated from one-tissue-compartment-models from a previous [$^{11}$C]UCB-J PET study in healthy individuals [19].

#### Functional magnetic resonance imaging analysis

##### *fMRI subsample and missing data*

Two participants (N-CT: *n*=1; N-ST: *n*=1) were not scanned due to suspected metal implants after enrolment in the trial. Moreover, baseline functional magnetic resonance imaging (fMRI) data was not available for two participants in the N-ST group due to technical issues with the screen used for viewing the fMRI tasks. Finally, seven individual scans (baseline: H-ST: *n*=3, N-ST: *n*=1; follow-up: H-CT: *n*=2, H-ST: *n*=1) were excluded from analysis due to the a priori defined movement exclusion criteria (see below). The remaining fMRI data was included in the linear mixed model analysis with the co-secondary outcome (i.e., mean percent BOLD signal change in the right DLPFC ROI). For the mechanistic and exploratory dorsal prefrontal cortex volume of interest and whole brain analyses conducted in FEAT, only the subjects with complete baseline and follow-up data were included (i.e., H-CT: *n*=31, H-ST: *n*=24, N-CT: *n*=27, and N-ST: *n*=24). The subsample with complete baseline and follow-up fMRI data (n=106) did not significantly differ from the remaining participants without complete fMRI data (n=20) on age, sex, verbal IQ, or years of education (*ps*≥0.34).

##### *Spatial working memory N-back task*

The functional magnetic resonance imaging (fMRI) paradigm was a spatial working memory N-back task that was programmed in E-prime 2.0 (Psychological Software Tools, Pittsburgh, USA) and shown on a screen that participants viewed through an angled mirror in the scanner. A yellow circle would appear randomly in a 5×5 grid for 300 ms followed by an empty grid for 1200 ms. Participants were instructed to indicate with a finger button press on a response pad whenever the yellow circle appeared in the same grid square as the one from *N* (1 or 2) steps back in the sequence (**Figure S4**). The task thus had two levels of cognitive load (i.e., 1-back and 2-back). Additionally, participants completed a sensorimotor control task (0-back), where they had to respond whenever the yellow circle appeared in any of the grid corners. Each condition block had 15 trials (with three targets) and were presented successively five times and interleaved by a fixation cross shown for eight seconds.

**Figure S4:** The visuospatial N-back working memory fMRI task. Participants viewed a grid square and were instructed to press a button on a response pad whenever the yellow circle appeared in the same grid square as the one shown *N* (1 or 2) steps back. This shows an example of the 1-back condition.

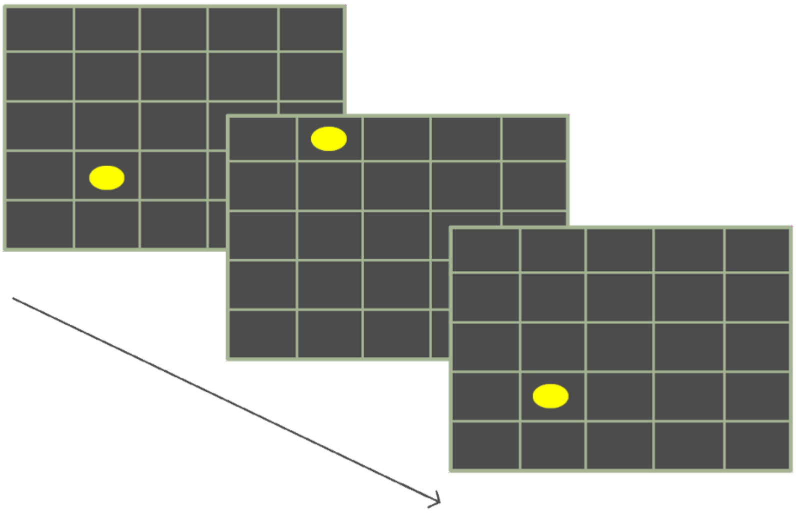

***Data acquisition***

Functional MRI data were acquired at the University Hospital of Copenhagen, Rigshospitalet, Denmark using a 3 Tesla SIEMENS Prisma scanner and a 32-channel head-neck coil. We obtained functional BOLD T2*2-weighted images with a multi-band multi-echo (MB3ME3) sequence (TR=1300 ms, TEs=15, 34, and 54 ms, flip angle=70°, matrix size=76x76, slices per volume=48, slice thickness=2.5 mm, voxel size=2.5 mm isotropic, and a 3x multiband acceleration factor). A total of 228 volumes were acquired for the spatial N-back task. We also acquired structural T1-weighted images to aid the registration of the functional images to a standard Montreal Neurological Institute (MNI) template using a standard Magnetization Prepared Rapid Gradient Echo (MPRAGE) pulse sequence (TR=2000 ms; TE=2.58 ms; flip angle=8°; matrix size=256x256; slices per volume=224; slice thickness=0.9 mm, and voxel size=0.9 mm isotropic). Finally, fieldmap data were acquired with a gradient echo sequence (TR=444 ms; TE1=4.92 ms; TE2=7.38 ms; flip angle=60°) to correct for geometric distortions due to $B_0$ inhomogeneities. A fieldmap image was computed based on the phase difference between two echoes using the FMRIB's Software Library (FSL) version 6.0.5.2 [20].

***Pre-processing***

The fMRI data were pre-processed using the fMRIPrep version 25.1.1 software package [21] and included generation of a BOLD reference volume by aligning and averaging three single-band references, estimation of head-motion parameters relative to the BOLD reference, correction for susceptibility distortion using the acquired field-map, slice-time correction, combination of the multi-echo times series, co-registration to the T-weighted image, resampling to the standard MNI152NLin6Asym space with a voxel size of 2x2x2, spatial smoothing using a five mm full-width-half-maximum Gaussian kernel, and high-pass temporal filtering using 100 seconds cut-off. We visually inspected each subject's registration to the MNI template and head movement parameters plots.

##### *First- and second-level analysis*

At the subject level, the task was modelled in the FMRI Expert Analysis Tool (FEAT) using two events: 2-back>0-back (general working memory) and 2-back>1-back (high-load working memory). The events were convolved with a double-gamma hemodynamic response function. We applied motion correction in the first-level general linear model (GLM) by including the six motion parameters estimated during motion correction as nuisance regressors. Framewise displacement (FD) was calculated as the sum of the absolute values of the derivative of the six motion parameters [22], and, for each time point, volumes with an FD>0.5 mm were identified and removed. We a priori decided to exclude subjects with a mean FD>0.3 mm *or* if the number of excluded volumes (due to movement) comprised more than 25% of the whole task (i.e., >57 of in total 228 volumes). Additional nuisance regressors were generated for these high-motion volumes with a value of 1 for the affected volume and 0 for all others. These regressors were included in the first-level analysis to account for motion-related artifacts through volume censoring (spike regression).

To investigate the effects of hypoxia and/or cognitive training on neural activity in other areas of the dorsal prefrontal cortex, we performed separate two-way mixed effects analyses of variance (ANOVA) in FEAT (i.e., comparing each H-CT, H-ST, N-CT group to the N-ST group). The dorsal prefrontal cortex volume of interest was defined on the standard Montreal Neurological Institute (MNI) template based on the Harvard-Oxford Cortical Structural Atlas probabilistic map thresholded at 5% [23] and included: the bilateral superior and medial frontal gyri, the frontal poles, and the superior parts of the anterior division of the cingulate gyrus. To explore any unpredicted treatment-related neural changes, we conducted similar analyses across the whole brain. All models included the two contrasts of interest and used FMRIB's Local Analysis of Mixed Effects (FLAME) as estimation method [24]. The significance level was set at $p<0.05$ and corrected for multiple comparisons with a cluster-forming threshold of $Z=2.56$ ($p<0.005$).

***Statistical analysis of in-scanner behavioural data***

In-scanner behavioural task performance was estimated using signal detection analysis by calculating the discriminability index (*d'*) using the following calculation: ((no. of hits + 0.5) / (total no. of targets + 1)) - ((no. of false alarms + 0.5) / (no. of distractors + 1)) [25]. The *d'* value was average for the 1-back and 2-back conditions. The *d'* score was missing at baseline for H-CT: $n$=1, N-CT: $n$=2, and N-ST: $n$=3 and at follow-up for H-CT: $n$=4, H-ST: $n$=3, N-CT: $n$=3, and N-ST: $n$=5. To investigate treatment effects on in-scanner task performance (i.e., *d'* score), we conducted linear mixed effects models with an unstructured covariance pattern. Fixed effects were time (baseline, treatment completion, and one-month follow-up), treatment (H-CT vs. H-ST vs. N-CT vs- N-ST), and their interaction (time*treatment). N-ST was set as reference group, and baseline constraint was applied.

**Treatment fidelity**

We examined treatment fidelity amongst the 114 participants who completed the full three-week intervention (H-CT: $n$=33, H-ST: $n$=27; N-CT: $n$=27, N-ST: $n$=27). Specifically, we monitored time spent outside the altitude treatment room during sessions (e.g., for the toilet break), as well as distance and time spent walking on the in-room treadmill. For the cognitive training, we calculated the total hours spent on training across sessions in addition to the maximum achieved levels across exercises that a participant had completed using data available from the HappyNeuron Pro. Finally, after completion of final one-month follow-up assessment and prior to unblinding, participants were asked to rate how much their treatment improved cognition in their daily lives. Participants rated this subjective treatment effect on a scale from 0-10, with 0 being no improvement, 5 being moderate, and 10 reflecting extreme improvement.

Group comparisons for normally distributed variables were assessed with one-way analyses of variance (ANOVAs) and, if significant, followed up with pairwise comparisons using Tukey's method for multiple comparisons adjustments. Group comparisons for nonparametric variables were performed with Kruskal-Wallis tests with Benjamini-Hochberg-adjusted pairwise comparisons. Effect sizes were computed as Eta-squared ($\eta^2$), where 0.01 were interpreted as small, 0.06 as medium, and >0.14 as large effects. Specifically for the maximum achieved levels across exercises, we compared only the two groups with the active cognitive training interventions (i.e., H-CT and N-CT) using independent samples $t$-tests with Cohen's $d$ as effect size; where 0.2 were interpreted as small, between 0.5 as medium, and ≥0.8 were interpreted as large effects. The results are presented in **Table S5**.

#### Treatment tolerability

##### *Procedure and measures*

To assess treatment tolerability, we collected the average $SpO_2$ and pulse rate for each treatment session (days 1-18) using data from the pulse oximeters. Analyses on daily average $SpO_2$ over treatment days 1-18 are available for a subsample of participants in [26]. Subjective altitude-related cerebral side effects were measured through the ESQ immediately after completion of each treatment session. The ESQ consists of 11 items, assessing symptoms like dizziness, headache, nausea, and others. Each item is rated on a scale from 0 to 5, where 0 indicates no experience of symptom, and 5 indicates experience to an extreme degree. The total score reflects the overall severity, ranging from a total of 0 to 55 points. We previously assessed the daily average ESQ total scores over days 1-18 as well as the percentage of moderate-to-extreme side effects (i.e., scores 3-5 on an item) in a subsample of participants – see [26] for further details. Participants were further interviewed about their daily subjective sleep quality the night before each treatment session (rated from 0-10) and subjective tiredness after each session (rated from 0-5), in addition to change in physical wellbeing from before to after a treatment session (rated from 0-10). Finally, all participants completed a VAS questionnaire assessing their subjective state (rated from 0-10) before and after each session.

##### *Comparisons of hypoxia versus normoxia*

We compared treatment tolerability for all participants who commenced their intervention and thereby had available data from at least one treatment day (incl. study dropouts) grouped into hypoxia ($n$=65) versus normoxia ($n$=57) arms. Group comparisons were assessed with independent samples $t$-tests and Mann-Whitney $U$ tests where appropriate. Effect sizes were determined with the rank-biserial correlation ($r$) for Mann-Whitney $U$ tests; where <0.3 were interpreted as small, between 0.3 to <0.5 as medium, and ≥0.5 were interpreted as large effects, or Cohen's $d$ for $t$-tests (see interpretation guidelines above). The results are reported in **Table S6**. In general, hypoxia (vs. normoxia) treatment was associated with average lower SpO2, higher pulse rate, higher ESQ total scores, increased subjective tiredness during sessions, increased feelings of dizziness from before to after sessions, and decreased subjective physical wellbeing from before to after sessions. However, effect sizes were or medium for all treatment tolerability measures, except for the lower SpO2 and higher pulse rate in hypoxia vs. normoxia groups.

***Comparisons of non-completers versus completers***

Finally, we compared treatment safety measures between participants who discontinued ($n$=8) versus completed their treatment ($n$=114) across both oxygen conditions. Comparisons were assessed with independent samples $t$-tests and Mann-Whitney $U$ tests where appropriate. Effect sizes were determined with the rank-biserial correlation ($r$) for Mann-Whitney $U$ tests or Cohen's $d$ for $t$-tests. The results are reported in **Table S7**. Across both oxygen/cognitive training conditions, participants who discontinued treatment were characterized by higher ESQ total scores in addition to increased subjective tiredness, sadness, dizziness, nausea, and decreased wellbeing from prior to after sessions. However, all of these differences were associated with small effect sizes, with the exception of the increased subjective tiredness in non-completers vs. completers.

**Biochemistry analysis**

***Haematological parameters***

Routine blood samples collected at baseline, prior to beginning days 8 and 19 of treatment, and follow-up were immediately analyzed for hemoglobin, erythrocytes, reticulocytes, and thrombocytes values by the Clinical Biochemistry Department, Section for External Projects, Copenhagen University Hospital. To assess the effects of hypoxia on hematological safety parameters, we grouped participants into hypoxia vs. normoxia groups and conducted linear mixed effect models. Fixed effects were time (baseline, day 8, day 19, and follow-up), treatment (hypoxia vs. normoxia), and their interaction (time*treatment). Normoxia was entered as the reference group, and baseline constraint was applied. All participants with baseline data were included in the analyses (hypoxia: $n$=66; normoxia: $n$=60). The raw mean and standard deviation values from the routine blood samples, missing data, and $p$-values estimated from the linear mixed effect models are reported in **Table S8** and visualised in **Figure S5**.

***Peripheral biomarkers***

Additional biomarker blood samples were collected at baseline (i.e., immediately prior to day 1 of treatment) and immediately prior to day 19 of treatment following a six-hour fasting period and were transferred to Frederiksberg Hospital and stored at -80 $^{o}$C until analysis. Serum concentrations of EPO (U-PLEX, #K151VXK-2), VEGF (V-PLEX, #K151RHD-2), and BDNF (U-PLEX, #K151WK-2) were quantified using the Meso Scale Discovery (MSD) electrochemiluminescence immunoassay platform. Measurements were performed on a SECTOR Imager 6000 plate reader according to the manufacturer's instructions (Meso Scale Diagnostics, Rockville, MD, USA). Data acquisition and analysis were conducted using MSD Workbench software. All samples were analysed in duplicate, and results are presented as mean values of the replicates. To investigate effects of hypoxia and/or cognitive training on peripheral biomarkers, we conducted linear mixed models with an unstructured covariance pattern. Fixed effects were time (baseline, treatment completion, and follow-up), treatment (H-CT vs. H-ST vs. N-CT vs. N-ST), and their interaction (time*treatment). N-ST was entered as the reference group, and baseline constraint was applied. The treatment effects, their 95% conficence intervals, and *p*-values are reported in **Table S10**.

To increase statistical power, we exploratorily combined hypoxia (*n*=65) vs. normoxia (*n*=57) and cognitive training (*n*=64) vs. sham training (*n*=58) groups and repeated the linear mixed models with each peripheral biomarker as outcome, respectively. Treatment effects, their 95% confidence intervals, and *p*-values from the linear mixed models are reported in **Table S11** and statistically significant results are visualised in **Figure S6**.

Repeated exposure to fixed 12% $O_2$ over three weeks may introduce acclimatization, which could alter the hypoxic dose [27]. To verify that possible group differences across haematological and peripheral biomarker measures reflected actual effects of hypoxia vs. normoxia, we conducted a post hoc analysis investigating $SpO_2$ values specifically at days 8 and 19 (i.e., days of blood sampling during treatment). Average $SpO_2$ values for hypoxia vs. normoxia groups were compared using Mann-Whitney *U* tests. Effect sizes were estimated using the rank-biserial correlation (*r*) (see interpretation above). The results are reported further below in the **Supplementary Results** (p. 35).

#### RESULTS

**Table S4:** Results of hypoxia and/or cognitive training versus normoxia sham training (N-ST) on each individual predefined tertiary outcome.

| | | **Treatment completion (week 4)** | | | **Follow-up (week 8)** | | |
|---|---|---|---|---|---|---|---|
| | Group | Treatment effect | 95% CI | *p*-value (adj.) | Treatment effect | 95% CI | *p*-value (adj.) |
| **Cognition** | | | | | | | |
| *Processing speed* | | | | | | | |
| RBANS Coding | H-CT | -0.09 | -0.41; 0.22 | 0.56 | -0.03 | -0.40; 0.33 | 0.86 |
| | H-ST | -0.25 | -0.58; 0.08 | 0.13 | -0.21 | -0.59; 0.18 | 0.28 |
| | N-CT | 0.04 | -0.29; 0.37 | 0.79 | -0.09 | -0.47; 0.30 | 0.66 |
| | N-ST | - | - | - | - | - | - |
| Trail Making Test A | H-CT | 0.58 | 0.22; 0.95 | **0.002** (0.06) | 0.08 | -0.23; 0.38 | 0.62 |
| | H-ST | 0.53 | 0.14; 0.91 | **0.01** (0.22) | 0.06 | -0.26; 0.38 | 0.71 |
| | N-CT | 0.45 | 0.07; 0.84 | **0.02** (0.37) | 0.22 | -0.10; 0.54 | 0.18 |
| | N-ST | - | - | - | - | - | - |
| *Attention* | | | | | | | |
| RBANS Digit Span | H-CT | 0.10 | -0.35; 0.56 | 0.65 | 0.51 | -0.03; 1.04 | 0.06 |
| | H-ST | -0.13 | -0.61; 0.35 | 0.60 | 0.13 | -0.43; 0.69 | 0.65 |
| | N-CT | 0.10 | -0.37; 0.57 | 0.68 | 0.35 | -0.20; 0.90 | 0.21 |
| | N-ST | - | - | - | - | - | - |
| RVP accuracy | H-CT | 0.16 | -0.09; 0.40 | 0.21 | 0.17 | -0.07; 0.42 | 0.17 |
| | H-ST | 0.19 | -0.07; 0.44 | 0.15 | 0.12 | -0.14; 0.38 | 0.35 |
| | N-CT | 0.09 | -0.16; 0.35 | 0.48 | 0.14 | -0.12; 0.40 | 0.30 |
| | N-ST | - | - | - | - | - | - |
| RVP mean latency | H-CT | -0.03 | -0.35; 0.29 | 0.86 | -0.03 | -0.35; 0.28 | 0.84 |
| | H-ST | 0.12 | -0.22; 0.45 | 0.50 | -0.05 | -0.38; 0.28 | 0.77 |
| | N-CT | -0.07 | -0.41; 0.27 | 0.68 | -0.21 | -0.54; 0.12 | 0.22 |

| | | | | | | | |
|---|---|---|---|---|---|---|---|
| | N-ST | - | - | - | - | - | - |
| RVP false alarms | H-CT | 0.35 | -0.05; 0.74 | 0.09 | 0.35 | -0.05; 0.76 | 0.09 |
| | H-ST | 0.18 | -0.24; 0.60 | 0.39 | 0.16 | -0.26; 0.59 | 0.45 |
| | N-CT | 0.19 | -0.23; 0.61 | 0.37 | 0.15 | -0.28; 0.57 | 0.49 |
| | N-ST | - | - | - | - | - | - |
| RVP probability of hit | H-CT | 0.11 | -0.16; 0.38 | 0.41 | 0.13 | -0.15; 0.41 | 0.34 |
| | H-ST | 0.18 | -0.11; 0.46 | 0.22 | 0.12 | -0.18; 0.41 | 0.43 |
| | N-CT | 0.07 | -0.21; 0.35 | 0.62 | 0.13 | -0.16; 0.42 | 0.38 |
| | N-ST | - | - | - | - | - | - |
| *Working memory* | | | | | | | |
| WAIS-III Letter-number sequencing | H-CT | 0.09 | -0.29; 0.47 | 0.65 | 0.50 | 0.15; 0.85 | **0.006** (0.11) |
| | H-ST | 0.05 | -0.35; 0.45 | 0.81 | 0.44 | 0.07; 0.81 | **0.02** (0.37) |
| | N-CT | 0.52 | 0.12; 0.92 | **0.01** (0.28) | 0.67 | 0.30; 1.04 | **<0.001 (0.03)** |
| | N-ST | - | - | - | - | - | - |
| SWM between errors | H-CT | 0.06 | -0.31; 0.43 | 0.76 | 0.13 | -0.20; 0.46 | 0.44 |
| | H-ST | 0.01 | -0.37; 0.40 | 0.94 | -0.03 | -0.37; 0.32 | 0.87 |
| | N-CT | 0.09 | -0.29; 0.48 | 0.63 | -0.11 | -0.45; 0.24 | 0.54 |
| | N-ST | - | - | - | - | - | - |
| SWM strategy | H-CT | 0.02 | -0.36; 0.40 | 0.91 | -0.08 | -0.50; 0.34 | 0.70 |
| | H-ST | 0.23 | -0.17; 0.63 | 0.26 | 0.02 | -0.41; 0.45 | 0.93 |
| | N-CT | 0.12 | -0.28; 0.51 | 0.57 | -0.12 | -0.55; 0.32 | 0.59 |
| | N-ST | - | - | - | - | - | - |
| *Verbal learning and memory* | | | | | | | |
| RAVLT total recall list I-V | H-CT | 0.26 | -0.09; 0.60 | 0.14 | 0.26 | -0.17; 0.70 | 0.23 |
| | H-ST | 0.10 | -0.26; 0.46 | 0.59 | 0.10 | -0.35; 0.56 | 0.65 |
| | N-CT | 0.31 | -0.06; 0.67 | 0.10 | 0.44 | -0.02; 0.90 | 0.06 |
| | N-ST | - | - | - | - | - | - |

| | | | | | | | |
|---|---|---|---|---|---|---|---|
| RAVLT immediate recall | H-CT | 0.24 | -0.12; 0.60 | 0.19 | 0.15 | -0.24; 0.53 | 0.46 |
| | H-ST | 0.30 | -0.08; 0.67 | 0.13 | 0.11 | -0.30; 0.52 | 0.59 |
| | N-CT | 0.43 | 0.05; 0.81 | **0.03** (0.37) | 0.33 | -0.08; 0.73 | 0.11 |
| | N-ST | - | - | - | - | - | - |
| RAVLT delayed recall | H-CT | 0.15 | -0.17; 0.48 | 0.35 | 0.22 | -0.15; 0.58 | 0.24 |
| | H-ST | 0.12 | -0.22; 0.46 | 0.49 | 0.19 | -0.20; 0.57 | 0.34 |
| | N-CT | 0.33 | -0.01; 0.67 | 0.06 | 0.41 | 0.02; 0.79 | **0.04** (0.38) |
| | N-ST | - | - | - | - | - | - |
| *Executive functions* | | | | | | | |
| Trail Making Test B | H-CT | 0.23 | -0.15; 0.61 | 0.23 | 0.13 | -0.21; 0.48 | 0.44 |
| | H-ST | 0.11 | -0.29; 0.50 | 0.59 | 0.05 | -0.32; 0.41 | 0.80 |
| | N-CT | 0.16 | -0.24; 0.56 | 0.42 | -0.04 | -0.41; 0.32 | 0.82 |
| | N-ST | - | - | - | - | - | - |
| Verbal fluency letter S | H-CT | 0.04 | -0.30; 0.37 | 0.83 | 0.03 | -0.29; 0.36 | 0.85 |
| | H-ST | 0.02 | -0.34; 0.37 | 0.93 | 0.30 | -0.04; 0.64 | 0.08 |
| | N-CT | 0.25 | -0.10; 0.60 | 0.15 | 0.29 | -0.05; 0.63 | 0.10 |
| | N-ST | - | - | - | - | - | - |
| Verbal fluency letter D | H-CT | 0.23 | -0.16; 0.62 | 0.24 | 0.21 | -0.13; 0.56 | 0.23 |
| | H-ST | 0.35 | -0.06; 0.76 | 0.09 | 0.49 | 0.13; 0.86 | **0.01** (0.22) |
| | N-CT | 0.38 | -0.03; 0.79 | 0.07 | 0.13 | -0.24; 0.49 | 0.49 |
| | N-ST | - | - | - | - | - | - |
| OTS problems solved on first choice | H-CT | -0.04 | -0.43; 0.35 | 0.83 | 0.03 | -0.35; 0.41 | 0.87 |
| | H-ST | -0.01 | -0.42; 0.41 | 0.98 | -0.01 | -0.42; 0.39 | 0.95 |
| | N-CT | 0.09 | -0.31; 0.50 | 0.66 | -0.04 | -0.43; 0.36 | 0.85 |
| | N-ST | - | - | - | - | - | - |
| OTS mean latency to correct | H-CT | 0.55 | 0.24; 0.86 | **<0.001** (0.06) | 0.30 | 0.02; 0.58 | **0.03** (0.33) |
| | H-ST | 0.16 | -0.17; 0.49 | 0.33 | -0.07 | -0.36; 0.22 | 0.62 |

| | | | | | | | |
|---|---|---|---|---|---|---|---|
| | N-CT | 0.33 | 0.01; 0.66 | **0.047** (0.37) | 0.24 | -0.05; 0.53 | 0.11 |
| | N-ST | - | - | - | - | - | - |
| WCST perseverative errors[1] | H-CT | -0.03 | -0.28; 0.21 | 0.79 | 0.20 | -0.08; 0.47 | 0.17 |
| | H-ST | -0.03 | -0.29; 0.23 | 0.81 | 0.11 | -0.18; 0.40 | 0.46 |
| | N-CT | 0.04 | -0.21; 0.30 | 0.74 | 0.28 | -0.01; 0.57 | 0.06 |
| | N-ST | - | - | - | - | - | - |
| *Facial expression recognition* | | | | | | | |
| ERT hit rate across emotions[2] | H-CT | -0.10 | -0.39; 0.19 | 0.49 | -0.24 | -0.60; 0.12 | 0.19 |
| | H-ST | 0.07 | -0.23; 0.37 | 0.64 | 0.02 | -0.36; 0.40 | 0.91 |
| | N-CT | 0.09 | -0.22; 0.40 | 0.56 | -0.14 | -0.52; 0.26 | 0.49 |
| | N-ST | - | - | - | - | - | - |
| ERT median reaction time across emotions[2] | H-CT | -0.32 | -0.62; 0.03 | **0.03** (0.33) | -0.09 | -0.32; 0.14 | 0.43 |
| | H-ST | -0.09 | -0.40; 0.21 | 0.55 | 0.10 | -0.13; 0.33 | 0.40 |
| | N-CT | -0.03 | -0.35; 0.28 | 0.84 | 0.11 | -0.13; 0.35 | 0.38 |
| | N-ST | - | - | - | - | - | - |
| *CAVIR* | | | | | | | |
| CAVIR global composite | H-CT | - | - | - | -0.04 | -0.28; 0.20 | 0.73 |
| | H-ST | - | - | - | 0.09 | -0.16; 0.34 | 0.48 |
| | N-CT | - | - | - | 0.11 | -0.14; 0.36 | 0.38 |
| | N-ST | - | - | - | - | - | - |
| **Questionnaires** | | | | | | | |
| WHOQoL-BREF | H-CT | -0.23 | -0.57; 0.12 | 0.19 | -0.24 | -0.58; 0.10 | 0.16 |
| | H-ST | -0.10 | -0.46; 0.26 | 0.58 | -0.18 | -0.53; 0.18 | 0.33 |
| | N-CT | -0.25 | -0.61; 0.11 | 0.17 | -0.20 | -0.55; 0.16 | 0.28 |
| | N-ST | - | - | - | - | - | - |
| AQoL | H-CT | -0.06 | -0.30; 0.19 | 0.66 | -0.22 | -0.55; 0.11 | 0.18 |
| | H-ST | -0.06 | -0.32; 0.19 | 0.63 | -0.16 | -0.51; 0.18 | 0.35 |

| | | | | | | | |
|---|---|---|---|---|---|---|---|
| | N-CT | -0.04 | -0.30; 0.21 | 0.74 | -0.08 | -0.43; 0.26 | 0.63 |
| | N-ST | - | - | - | - | - | - |
| COBRA | H-CT | -0.10 | -0.37; 0.16 | 0.43 | -0.10 | -0.38; 0.18 | 0.50 |
| | H-ST | -0.22 | -0.50; 0.05 | 0.12 | -0.13 | -0.43; 0.16 | 0.37 |
| | N-CT | -0.09 | -0.36; 0.19 | 0.53 | -0.09 | -0.38; 0.20 | 0.54 |
| | N-ST | - | - | - | - | - | - |
| PSQI | H-CT | 0.15 | -0.20; 0.51 | 0.40 | -0.03 | -0.44; 0.38 | 0.88 |
| | H-ST | 0.09 | -0.28; 0.46 | 0.63 | 0.23 | -0.21; 0.66 | 0.30 |
| | N-CT | 0.03 | -0.35; 0.40 | 0.89 | -0.07 | -0.51; 0.36 | 0.74 |
| | N-ST | - | - | - | - | - | - |
| WSAS | H-CT | 0.08 | -0.20; 0.35 | 0.58 | -0.22 | -0.58; 0.14 | 0.23 |
| | H-ST | -0.19 | -0.48; 0.10 | 0.19 | -0.18 | -0.55; 0.20 | 0.35 |
| | N-CT | -0.20 | -0.48; 0.09 | 0.18 | -0.12 | -0.49; 0.26 | 0.54 |
| | N-ST | - | - | - | - | - | - |
| SDS | H-CT | -0.13 | -0.53; 0.27 | 0.53 | -0.18 | -0.55; 0.19 | 0.33 |
| | H-ST | -0.35 | -0.77; 0.08 | 0.11 | -0.25 | -0.64; 0.14 | 0.20 |
| | N-CT | 0.03 | -0.39; 0.45 | 0.89 | 0.01 | -0.38; 0.39 | 0.97 |
| | N-ST | - | - | - | - | - | - |

**Note:** All cognitive and questionnaire scores are *z*-standardized based on the whole sample mean and standard deviations at baseline. Includes all available data (incl. study dropouts). [1]Missing data at baseline for N-CT: *n*=1; missing data at treatment completion for H-ST: *n*=1; missing data at follow-up for H-CT: *n*=2 and N-ST: *n*=4.[2]Missing data at baseline for N-CT: *n*=4 and N-ST: *n*=2; missing data at treatment completion for H-CT: *n*=1, N-CT: n=3, and N-ST: n=2; missing data at follow-up for N-CT: n=3 and N-ST: n=2. **Abbreviations:** AQoL=Assessment of Quality of Life; CAVIR=Cognition Assessment in Virtual Reality; CI=Confidence interval; COBRA=Cognitive Complaints in Bipolar Disorder Rating Assessment; ERT=Emotion Recognition Test; H-CT=Hypoxia with cognitive training; H-ST=Hypoxia with sham training; N-CT=Normoxia with cognitive training; N-ST=Normoxia with sham training; OTS=One Touch Stockings of Cambridge; RAVLT=Rey Auditory Verbal Learning Test; RBANS=Repeatable Battery for the Assessment of Neuropsychological Status; PSQI=Pittsburgh Sleep Quality Index; RVP=Rapid Visual Information Processing Test; SDS=Sheehan Disability Scale; SWM=Spatial Working Memory; WAIS=Wechsler Adult Intelligence Scale; WCST=Wisconsin Card Sorting Test; WHOQol-BREF=World Health Organization Quality of Life; WSAS=Work and Social Adjustment Scale.

**Table S5:** Treatment fidelity comparisons among the four treatment groups for participants who completed the three-week intervention.

| | **H-CT** | **H-ST** | **N-CT** | **N-ST** | **Effect size** | ***p*-value** |
|---|---|---|---|---|---|---|
| *N* | 33 | 27 | 27 | 27 | | |
| No. treatment sessions | 17.0 (16.0-18.0) | 17.0 (16.0-18.0) | 17.0 (17.0-18.0) | 17.0 (16.0-18.0) | 0.004 | 0.33 |
| Subjective treatment effect (0-10)[1] | 6.0 (4.0-6.0) | 1.0 (0.0-4.0) | 5.0 (2.5-6.0) | 2.0 (0.0-4.0) | 0.25 | **<0.001[2]** |
| Hours spent on cognitive/sham training | 30.8 (5.8) | 26.6 (4.1) | 31.1 (6.0) | 27.9 (4.1) | 0.12 | **0.002[3]** |
| Average highest task level achieved | 12.7 (3.3) | - | 14.7 (3.2) | - | -0.64 | **0.02** |
| Time on treadmill, minutes | 25.6 (23.9-26.5) | 25.2 (23.8-27.1) | 26.4 (24.7-27.1) | 26.0 (24.7-27.8) | -0.005 | 0.49 |
| Distance on treadmill, kilometers | 1.2 (1.1-1.4) | 1.1 (0.9-1.3) | 1.2 (1.0-1.3) | 1.2 (1.0-1.3) | -0.008 | 0.54 |
| Minutes spent outside treatment room | 1.6 (0.4-2.9) | 0.7 (0.4-1.4) | 0.0 (0.0-0.6) | 0.3 (0.0-0.7) | 0.21 | **<0.001[4]** |

**Note:** Continuous values are shown as either "n (n)" (reflecting means and standard deviations) or "n (n-n)" (indicating median and its 25th and 75th percentiles). Effect sizes are computed as Eta-squared (η2) comparisons across all four arms and Cohen's *d* for comparisons between the two training groups. Includes data for all participants who completed the three-week intervention (*n*=114). [1]Missing data for H-CT: *n*=2, N-CT: *n*=3, and N-ST: *n*=1. [2]Differences between the two groups with cognitive training vs. the two groups with sham training ($ps \leq 0.01$). [3]Differences between H-ST vs. the two groups with cognitive training ($ps \leq 0.01$). [4]Differences between the two hypoxia groups vs. the normoxia groups ($ps \leq 0.02$). **Abbreviations:** H-CT=Hypoxia with cognitive training; H-ST=Hypoxia with sham training; N-CT=Normoxia with cognitive training; N-ST=Normoxia with sham training.

**Table S6:** Treatment tolerability comparisons for all participants who commenced their treatment grouped into hypoxia versus normoxia treatment groups.

| | **Hypoxia** | **Normoxia** | **Effect size** | ***p*-value** |
|---|---|---|---|---|
| *N* | 65 | 57 | | |
| SpO2 | 86.4 (84.7-87.6) | 97.5 (97.1-97.8) | 0.86 | **<0.001** |
| Pulse | 81.5 (8.5) | 75.8 (7.3) | 0.71 | **<0.001** |
| ESQ total score (0-55) | 2.7 (1.6-5.2) | 1.2 (0.5-3.1) | 0.31 | **<0.001** |
| Subjective sleep quality (0-10) | 8.6 (8.2-9.2) | 8.7 (8.2-9.1) | <0.01 | 0.95 |
| Subjective tiredness (0-5) | 2.1 (0.8) | 1.7 (0.9) | 0.44 | **0.02** |
| Change in physical wellbeing | -0.2 (-0.5-0.0) | 0.0 (-0.2-0.1) | 0.30 | **0.001** |
| VAS change in happiness | -0.1 (-0.3-0.0) | -0.1 (-0.5-0.0) | 0.08 | 0.37 |
| VAS change in wellbeing | -0.2 (-0.5-0.0) | -0.1 (-0.5-0.0) | 0.05 | 0.62 |
| VAS change in sadness | 0.0 (-0.1-0.1) | 0.0 (-0.2-0.3) | 0.02 | 0.87 |
| VAS change in vigilance | -0.2 (-0.6-0.0) | -0.2 (-0.6-0.0) | 0.04 | 0.70 |
| VAS change in anxiety | 0.0 (-0.1-0.0) | 0.0 (0.0-0.1) | 0.17 | 0.07 |
| VAS change in dizziness | 0.4 (0.1-0.9) | 0.0 (0.0-0.3) | 0.39 | **<0.001** |
| VAS change in nausea | 0.1 (0.0-0.2) | 0.0 (-0.1-0.2) | 0.13 | 0.17 |

**Note:** Continuous values are shown as either "n (n)" (reflecting means and standard deviations) or "n (n-n)" (indicating median and its 25th and 75th percentiles). Effect sizes were determined with the rank-biserial correlation (*r*) for Mann-Whitney *U* tests or Cohen's *d* for *t*-tests. Includes all available data (incl. study dropouts). All VAS items are rated on a scale from 0-10, with higher scores indicating greater presence of a feeling/state. Positive VAS change scores thus indicate a higher presence of a feeling/state from prior to immediately after a treatment session and vice versa. **Abbreviations:** ESQ=Environmental Symptoms Questionnaire; VAS=Visual Analogue Scale.

**Table S7:** Treatment tolerability comparisons for participants who discontinued their treatment compared to participants who completed their treatment across both oxygen/training conditions.

| | **Non-completers** | **Completers** | **Effect size** | ***p*-value** |
|---|---|---|---|---|
| *N* | 8 | 114 | | |
| SpO2 | 89.0 (87.7-97.1) | 89.2 (85.7-97.5) | <0.01 | 0.94 |
| Pulse | 77.2 (9.2) | 79.0 (8.4) | 0.21 | 0.57 |
| ESQ total score (0-55) | 12.5 (4.7-14.2) | 1.8 (0.8-4.0) | 0.30 | **0.001** |
| Subjective sleep quality (0-10) | 8.2 (7.5-8.3) | 8.6 (8.3-9.1) | 0.19 | 0.08 |
| Subjective tiredness (0-5) | 2.5 (0.6) | 1.9 (0.8) | 0.80 | **0.03** |
| Change in physical wellbeing | -0.5 (-1.7-0.0) | -0.1 (-0.3-0.1) | 0.19 | **0.04** |
| VAS change in happiness | -0.3 (-1.6-0.0) | -0.1 (-0.5-0.1) | 0.15 | 0.11 |
| VAS change in wellbeing | -0.6 (-2.0--0.2) | -0.2 (-0.5-0.0) | 0.22 | **0.02** |
| VAS change in sadness | 0.3 (0.0-2.2) | 0.0 (-0.2-0.2) | 0.20 | **0.03** |
| VAS change in vigilance | -0.8 (-1.1-0.0) | -0.2 (-0.6-0.0) | 0.14 | 0.14 |
| VAS change in anxiety | 0.0 (-0.1-0.0) | 0.0 (-0.1-0.0) | 0.08 | 0.37 |
| VAS change in dizziness | 1.7 (1.3-2.4) | 0.1 (0.0-0.5) | 0.35 | **<0.001** |
| VAS change in nausea | 0.6 (0.4-2.3) | 0.0 (-0.1-0.2) | 0.33 | **<0.001** |

**Note:** Continuous values are shown as either "n (n)" (reflecting means and standard deviations) or "n (n-n)" (indicating median and its 25th and 75th percentiles). Effect sizes were determined with the rank-biserial correlation (*r*) for Mann-Whitney *U* tests or Cohen's *d* for *t*-tests. Includes all available data (incl. study dropouts). All VAS items are rated on a scale from 0-10, with higher scores indicating greater presence of a feeling/state. Positive VAS change scores thus indicate a higher presence of a feeling/state from prior to immediately after a treatment session and vice versa. **Abbreviations:** ESQ=Environmental Symptoms Questionnaire; VAS=Visual Analogue Scale.

**Table S8:** Haematological safety parameters mean raw scores and standard deviations, in addition to *p*-values from the linear mixed model analyses for all participants grouped into hypoxia versus normoxia treatments at baseline, day 8, day 19, and at one-month follow-up.

| | **Hypoxia** | **Normoxia** | ***p*-value** |
|---|---|---|---|
| *N* | 66 | 60 | |
| Reticulocytes baseline, mean (SD) | 13.3 (5.0) | 13.8 (3.9) | - |
| Reticulocytes day 8, mean (SD) | 16.0 (4.9) | 14.4 (3.5) | **0.01** |
| Reticulocytes day 19, mean (SD) | 14.3 (4.4) | 14.1 (3.2) | 0.19 |
| Reticulocytes follow-up, mean (SD) | 14.5 (5.4) | 13.9 (3.9) | 0.38 |
| Erythrocytes baseline, mean (SD) | 0.4 (0.0) | 0.4 (0.0) | - |
| Erythrocytes day 8, mean (SD) | 0.4 (0.0) | 0.4 (0.0) | 0.11 |
| Erythrocytes day 19, mean (SD) | 0.4 (0.0) | 0.4 (0.0) | **0.02** |
| Erythrocytes follow-up, mean (SD) | 0.4 (0.0) | 0.4 (0.0) | 0.75 |
| Haemoglobin baseline, mean (SD) | 8.8 (0.7) | 8.8 (0.7) | - |
| Haemoglobin day 8, mean (SD) | 9.0 (0.8) | 8.8 (0.7) | **0.01** |
| Haemoglobin day 19, mean (SD) | 9.1 (0.8) | 8.7 (0.6) | **<0.001** |
| Haemoglobin follow-up, mean (SD) | 8.9 (0.8) | 8.8 (0.6) | 0.98 |
| Thrombocytes baseline, mean (SD) | 247.3 (49.1) | 254.8 (59.1) | - |
| Thrombocytes day 8, mean (SD) | 269.0 (52.2) | 266.5 (54.9) | **0.04** |
| Thrombocytes day 19, mean (SD) | 271.5 (51.2) | 249.8 (48.4) | **<0.001** |
| Thrombocytes follow-up, mean (SD) | 247.1 (64.9) | 251.9 (49.1) | 0.71 |

**Note:** Values are shown as raw means and standard deviations. Includes all available data (incl. study dropouts). Missing data at day 8 of treatment for hypoxia: $n$=19 and normoxia: $n$=15; missing data at day 19 of treatment for hypoxia: $n$=9 and normoxia: $n$=10; missing data at follow-up for hypoxia: $n$=28 and normoxia: $n$=26. **Abbreviations:** SD=standard deviation.

**Figure S5:** Treatment effects on haematological parameters for hypoxia versus normoxia. Values are shown in percent (mean for each group for each time point). Error bars show the standard error of the mean (s.e.m.). Haematological parameter changes and comparisons. *$p$<0.05 according to linear mixed models based on raw values.

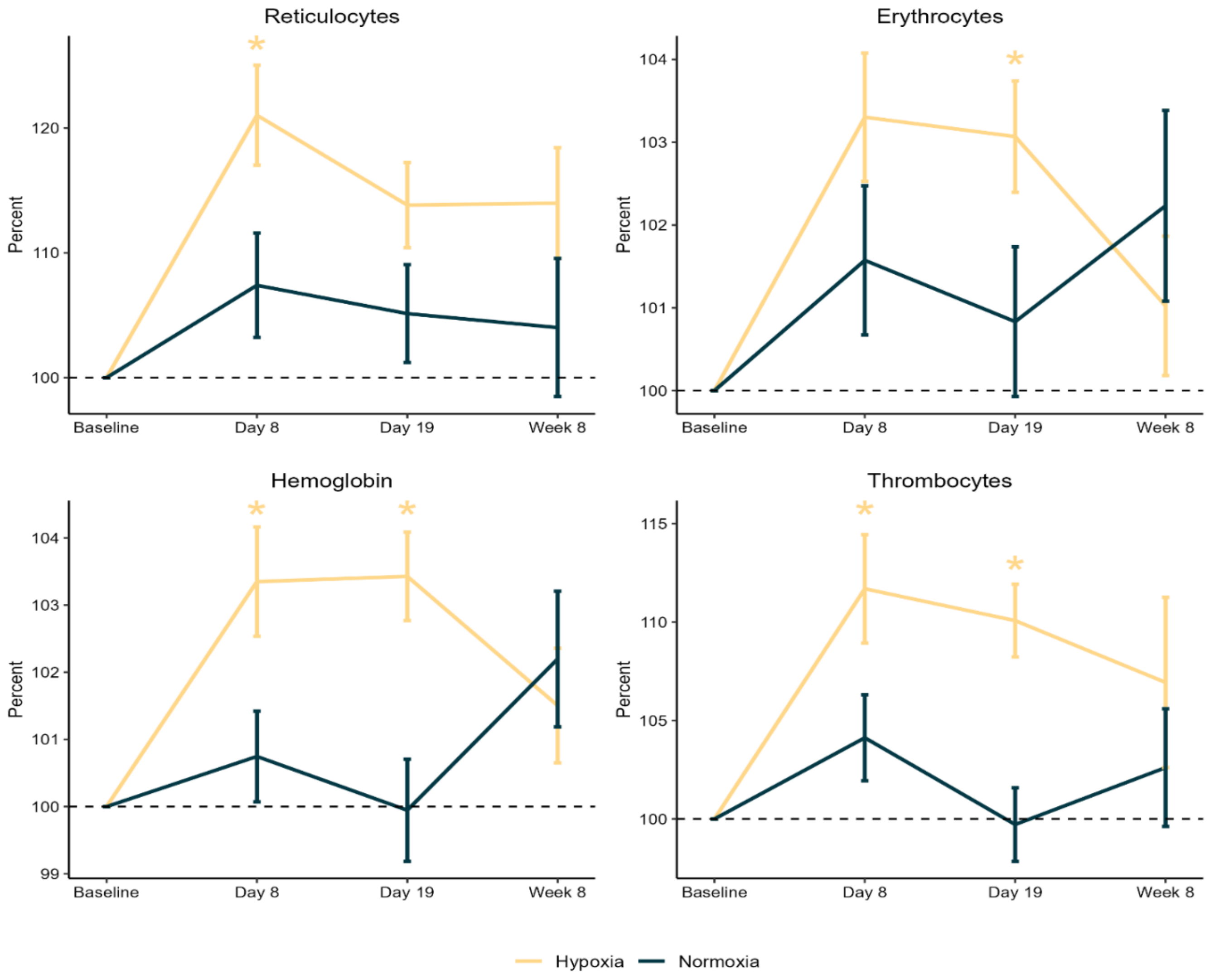

**Table S9**. Group comparisons of average [$^{11}$C]UCB-J synaptic vesicle glycoprotein 2A (SV2A) binding in the additional exploratory regions of interest (ROIs) at treatment completion.

| | **H-CT** | **N-ST** | **Cohen's *d*** | ***p*-value** |
|---|---|---|---|---|
| *N* | 16 | 14 | | |
| Neocortex | 3.59 (0.37) | 3.78 (0.24) | -0.57 | 0.12 |
| Anterior cingulate cortex | 3.80 (0.48) | 4.00 (0.36) | -0.46 | 0.21 |
| Posterior cingulate cortex | 3.95 (0.76) | 4.35 (0.42) | -0.64 | 0.08 |
| Gyrus rectus | 3.47 (0.93) | 3.99 (0.25) | -0.75 | **0.04** |
| Orbitofrontal gyri | 3.46 (0.46) | 3.75 (0.24) | -0.79 | **0.03** |
| Inferior frontal gyrus | 3.92 (0.41) | 4.04 (0.23) | -0.35 | 0.33 |
| Superior frontal gyrus | 3.60 (0.34) | 3.69 (0.35) | -0.28 | 0.44 |
| Middle frontal gyrus | 3.80 (0.34) | 3.89 (0.32) | -0.29 | 0.43 |
| Precentral gyrus | 3.48 (0.34) | 3.62 (0.32) | -0.43 | 0.25 |
| Postcentral gyrus | 3.41 (0.31) | 3.54 (0.24) | -0.48 | 0.20 |
| Superior parietal gyrus | 3.57 (0.35) | 3.76 (0.26) | -0.60 | 0.10 |
| Inferolateral remainder of parietal lobe | 3.76 (0.36) | 3.95 (0.24) | -0.62 | 0.09 |
| Anterior temporal lobe (medial) | 3.12 (0.44) | 3.28 (0.24) | -0.43 | 0.23 |
| Anterior temporal lobe (lateral) | 3.71 (0.49) | 3.91 (0.25) | -0.49 | 0.18 |
| Posterior temporal lobe | 3.61 (0.44) | 3.84 (0.26) | -0.61 | 0.10 |
| Parahippocampal and ambient gyri | 2.79 (0.40) | 2.96 (0.25) | -0.48 | 0.18 |
| Superior temporal gyrus | 3.81 (0.48) | 3.99 (0.20) | -0.47 | 0.20 |
| Middle and inferior temporal gyri | 3.84 (0.48) | 4.02 (0.24) | -0.47 | 0.20 |
| Fusiform gyrus | 3.44 (0.35) | 3.55 (0.27) | -0.36 | 0.32 |
| Lingual gyrus | 3.63 (0.42) | 3.92 (0.31) | -0.76 | **0.04** |
| Cuneus | 3.65 (0.46) | 3.96 (0.33) | -0.77 | **0.04** |
| Lateral remainder of occipital lobe | 3.38 (0.41) | 3.61 (0.28) | -0.66 | 0.08 |
| Insula | 3.55 (0.47) | 3.70 (0.19) | -0.41 | 0.26 |
| Amygdala | 3.27 (0.54) | 3.53 (0.25) | -0.61 | 0.09 |
| Nucleus accumbens | 4.33 (0.44) | 4.57 (0.33) | -0.60 | 0.11 |
| Caudate nucleus | 3.94 (0.53) | 4.12 (0.25) | -0.43 | 0.23 |
| Putamen | 4.55 (0.53) | 4.81 (0.22) | -0.61 | 0.09 |
| Thalamus | 3.43 (0.52) | 3.52 (0.15) | -0.24 | 0.49 |
| Striatum | 4.26 (0.52) | 4.47 (0.22) | -0.52 | 0.15 |

**Note:** Continuous values are shown as "n (n)" (reflecting means and standard deviations). All regions are bilateral. **Abbreviations:** H-CT=Hypoxia with cognitive training; N-ST=Normoxia with sham training.

**Table S10:** Effects of hypoxia and/or cognitive training versus normoxia sham training on blood serum levels of EPO, VEGF, and BDNF from baseline to day 19 of treatment.

| | | **Day 19 of treatment** | | |
|---|---|---|---|---|
| | Group | Treatment effect | 95% CI | *p*-value |
| Serum EPO | H-CT | -0.23 | -0.48; 0.32 | 0.09 |
| | H-ST | -0.24 | -0.51; 0.03 | 0.08 |
| | N-CT | 0.08 | -0.19; 0.35 | 0.54 |
| | N-ST | - | - | - |
| Serum VEGF | H-CT | 0.21 | -0.10; 0.51 | 0.18 |
| | H-ST | 0.26 | -0.05; 0.57 | 0.10 |
| | N-CT | 0.09 | -0.22; 0.40 | 0.55 |
| | N-ST | - | - | - |
| Serum BDNF | H-CT | -0.46 | -0.93; 0.02 | 0.06 |
| | H-ST | -0.26 | -0.76; 0.23 | 0.29 |
| | N-CT | -0.62 | -1.12; -0.13 | **0.01** |
| | N-ST | - | - | - |

**Note:** All blood-based biomarker values are *z*-standardized based on the whole sample's means and standard deviations at baseline. Includes all available data, but only for participants who commenced their treatment (incl. study dropouts). Missing data at baseline for H-CT: *n*=6, H-ST: *n*=1, and N-ST: *n*=1; missing data at day 19 of treatment for H-CT: *n*=1, H-ST: *n*=2, and N-CT: *n*=2. **Abbreviations:** BDNF=Brain-derived neurotrophic factor; CI=Confidence interval; EPO=Erythropoietin; H-CT=Hypoxia with cognitive training; H-ST=Hypoxia with sham training; N-CT=Normoxia with cognitive training; N-ST=Normoxia with sham training; VEGF=Vascular endothelial growth factor.

**Table S11:** Effects of (i) hypoxia versus normoxia treatment and (ii) cognitive versus sham training on serum levels of erythropoietin (EPO), vascular endothelial growth factor (VEGF), and brain-derived neurotrophic factor (BDNF) from baseline to day 19 of treatment.

| | | **Day 19 of treatment** | | |
|---|---|---|---|---|
| | Comparison | Treatment effect | 95% CI | *p*-value |
| Serum EPO | Hypoxia vs. normoxia | -0.25 | -0.42; -0.08 | **0.005** |
| | Cognitive vs. sham training | -0.02 | -0.20; 0.16 | 0.81 |
| Serum VEGF | Hypoxia vs. normoxia | 0.20 | 0.01; 0.39 | **0.04** |
| | Cognitive vs. sham training | 0.04 | -0.15; 0.24 | 0.66 |
| Serum BDNF | Hypoxia vs. normoxia | -0.06 | -0.39; 0.27 | 0.72 |
| | Cognitive vs. sham training | -0.42 | -0.75; -0.10 | **0.01** |

**Note:** All blood-based biomarker values are *z*-standardized based on the whole sample's means and standard deviations at baseline. Includes all available data, but only for participants who commenced their treatment (incl. study dropouts). Missing data at baseline for H-CT: *n*=6, H-ST: *n*=1, and N-ST: *n*=1; missing data at day 19 of treatment for H-CT: *n*=1, H-ST: *n*=2, and N-CT: *n*=2. **Abbreviations:** BDNF=Brain-derived neurotrophic factor; CI=Confidence interval; EPO=Erythropoietin; VEGF=Vascular endothelial growth factor.

**Figure S6:** Statistically significant effects of hypoxia (vs. normoxia) and cognitive training (vs. sham) groups on serum concentrations of erythropoietin (EPO), vascular endothelial growth factor (VEGF), or brain-derived neurotrophic factor (BDNF) from baseline (dotted line) to day 19 of treatment. Values are shown in percent (mean for each group). Error bars show the standard error of the mean (s.e.m.).

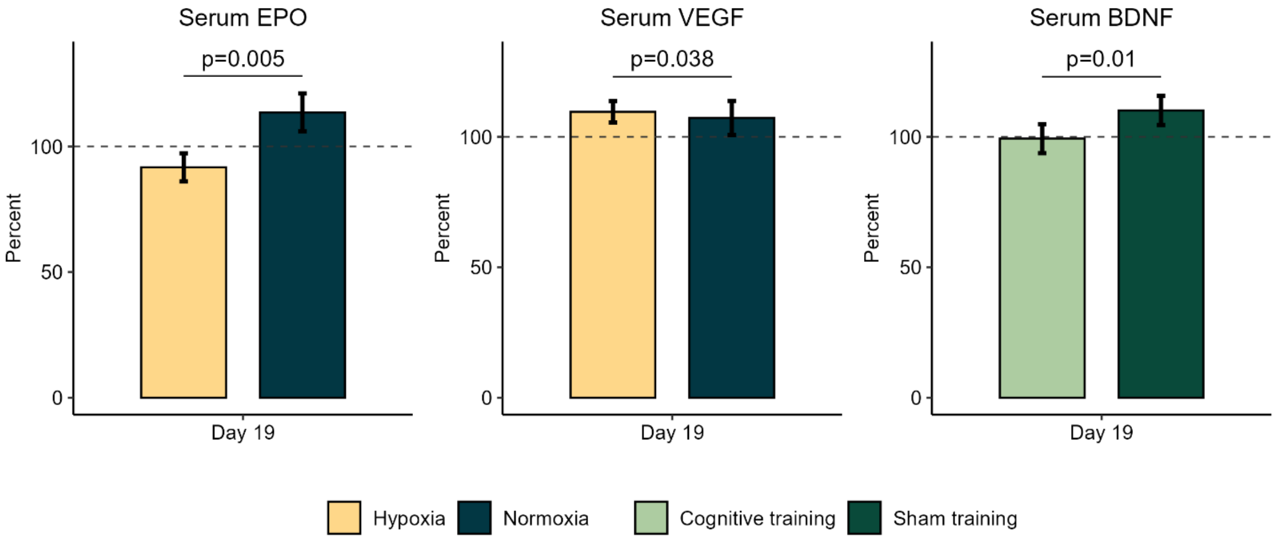

#### Post hoc analysis of $SpO_2$ on blood sampling days

Repeated exposure to 12% $O_2$ over three weeks may introduce acclimatization, which could alter the hypoxic dose [27]. To confirm that the observed group differences in both haematological and peripheral biomarkers indeed reflect *actual* hypoxia-related effects, we explored $SpO_2$ values specifically at days 8 and 19 (i.e., days of blood sampling during treatment). We compared average $SpO_2$ in hypoxia vs. normoxia groups using Mann-Whitney $U$ tests. At both treatment days, hypoxia was associated with reduced average $SpO_2$ values (day 8 - hypoxia: $Mdn$=86% (IQR 84.1-88.2); normoxia: $Mdn$=97.5% (IQR 96.7-98.1), $p$<0.001, $r$=0.86; day 19 - hypoxia: $Mdn$=86.5% (IQR 85.0-88.6); normoxia: $Mdn$=97.5% (IQR 97.0-98.0), $p$<0.001, $r$=0.86), which confirms that the observed haematological changes reflect a stable hypoxic stimulus.
